# Delayed Arousal Response to Sleep Apnea Encodes Mortality

**DOI:** 10.64898/2026.05.18.26353387

**Authors:** Jiahao Fan, M. Brandon Westover, Yue Leng, Guo-Qiang Zhang, Katie L Stone, Susan Redline, Robert J. Thomas, Licong Cui, Haoqi Sun

**Affiliations:** Department of Neurology, McGovern Medical School, The University of Texas Health Science Center at Houston, Houston, TX 77030, USA; Department of Neurology, Beth Israel Deaconess Medical Center, Harvard Medical School, Boston, MA 02215, USA; Department of Psychiatry and Behavioral Sciences, University of California, San Francisco, CA 94107, USA; McWilliams School of Biomedical Informatics, The University of Texas Health Science Center at Houston, Houston, TX 77030, United States; California Pacific Medical Center Research Institute, San Francisco, CA 94158, USA; Department of Epidemiology and Biostatistics, University of California, San Francisco, San Francisco, CA 94143, USA; Division of Sleep and Circadian Disorders, Department of Medicine, Brigham and Women’s Hospital, Boston, MA 02115, USA; Division of Sleep Medicine, Harvard Medical School, Boston, MA 02215, USA; Division of Pulmonary, Critical Care and Sleep Medicine, Department of Medicine, Beth Israel Deaconess Medical Center, Harvard Medical School, Boston, MA 02215, USA

**Keywords:** Sleep, Arousal, Obstructive Sleep Apnea, Mortality, Cardiovascular Diseases

## Abstract

**Rationale:** Conventional measures of obstructive sleep apnea severity, particularly the apnea-hypopnea index, do not adequately capture event-level neurophysiologic responses to respiratory events. Whether post-apnea/hypopnea arousal dynamics provide prognostic information beyond established metrics remains unknown.

**Objectives:** To determine whether post-apnea/hypopnea arousal dynamics are associated with all-cause and cardiovascular mortality.

**Methods:** We conducted a retrospective analysis of in-home polysomnography data from 8,053 adults across four community-based cohorts. Peak time (PT; latency to maximal arousal probability), peak height (PH; maximal arousal probability), and area under the curve (AUC; cumulative arousal probability) were derived from peri-stimulus time histograms aligned to event termination. Associations with mortality were examined using multivariable Cox models and random-effects meta-analysis.

**Measurements and Main Results:** PT, but not PH or AUC, was associated with mortality. In pooled analyses, each 1-second delay in PT was associated with higher all-cause mortality in males (hazard ratio [HR], 1.04; 95% confidence interval [CI], 1.02–1.06) and females (HR, 1.03; 95% CI, 1.00–1.06). For cardiovascular mortality, each 1-second delay in PT was associated with higher risk in males (HR, 1.05; 95% CI, 1.02–1.08) but not females (HR, 1.04; 95% CI, 0.99–1.10). Associations were driven primarily by non-rapid eye movement sleep and remained materially unchanged after additional adjustment for apnea-hypopnea index, arousal index, and hypoxic burden.

**Conclusions:** Delayed arousal timing after apnea/hypopnea termination was associated with increased mortality risk independent of conventional measures of obstructive sleep apnea severity. Event-level arousal timing may provide prognostic information beyond count-based and hypoxemia-based metrics.

## Introduction

Sleep involves multi-organ network physiology, requiring coordinated interactions among the brain, respiratory, autonomic, and cardiovascular systems. Sleep disorders impose stressful events on the network, such as sleep apnea, which contribute to a range of unfavorable health outcomes in adults (1–5), such as cardiovascular diseases (CVD), dementia (6), and mortality. However, conventional measures of obstructive sleep apnea (OSA) severity, particularly the apnea-hypopnea index (AHI), do not account for the temporal response to the apneic stressor at the event level. AHI correlates inconsistently with long-term health outcomes (5, 7, 8).

Several metrics have been proposed to go beyond the AHI. Hypoxic burden (HB), which integrates the depth and duration of oxygen desaturations time-aligned to apneic events, has shown a stronger association with CVD mortality than the AHI in community-based cohorts (9). Another metric, the apnea-induced heart rate response (ΔHR), which quantifies the surge in heart rate time-aligned to apneic events as an index of sympathetic activation, shows positive associations with risks of CVD and death (10). These measures highlight the importance of event-level physiological information.

Here, we used the stressor-response framework (11) based on the average temporal dynamics of the response, event-aligned to the stressor, represented as a peristimulus time histogram (PSTH) (12). In this study, we illustrate this framework using arousal as the response event and apnea as the stressor event across four large, community-based cohorts. We hypothesized that the temporal dynamics of post-apnea/hypopnea (post-stressor) arousals (response) are associated with the risk of all-cause mortality and CVD mortality.

## Methods

### Study design

This is a retrospective cohort study of adult participants from four community-dwelling cohorts: Sleep Heart Health Study (SHHS: 1995-2003) (13, 14), Multi-Ethnic Study of Atherosclerosis (MESA: 2010-2013) (15), Osteoporotic Fractures in Males (MrOS: 2003-2005) (16–18), and Study of Osteoporotic Fractures (SOF: 2002-2004) (19). All cohorts included overnight at-home polysomnography (PSG) performed by centrally trained technicians. All cohort committees approved the use of the data. Written consent was obtained in each cohort. The study protocol was also reviewed and approved by the Committee for the Protection of Human Subjects at the University of Texas Health Science Center at Houston (IRB# HSC-SBMI-25-0007). The study follows the Strengthening the Reporting of Observational Studies in Epidemiology (STROBE) (20) guideline.

The inclusion criteria were: 1) PSG recording time at least 5 hours; 2) availability of respiratory events (apnea and hypopnea) annotations and AHI greater than 5/hour; 3) post-apnea/hypopnea arousal detected within a pre-defined time window; 4) availability of follow-up data on all-cause and CVD mortality; and 5) availability of covariates.

Detailed information for all covariates across the cohorts are provided in the Supplement. Figure 1 presents a flow diagram detailing the inclusion criteria used for each cohort.

**Figure 1.**
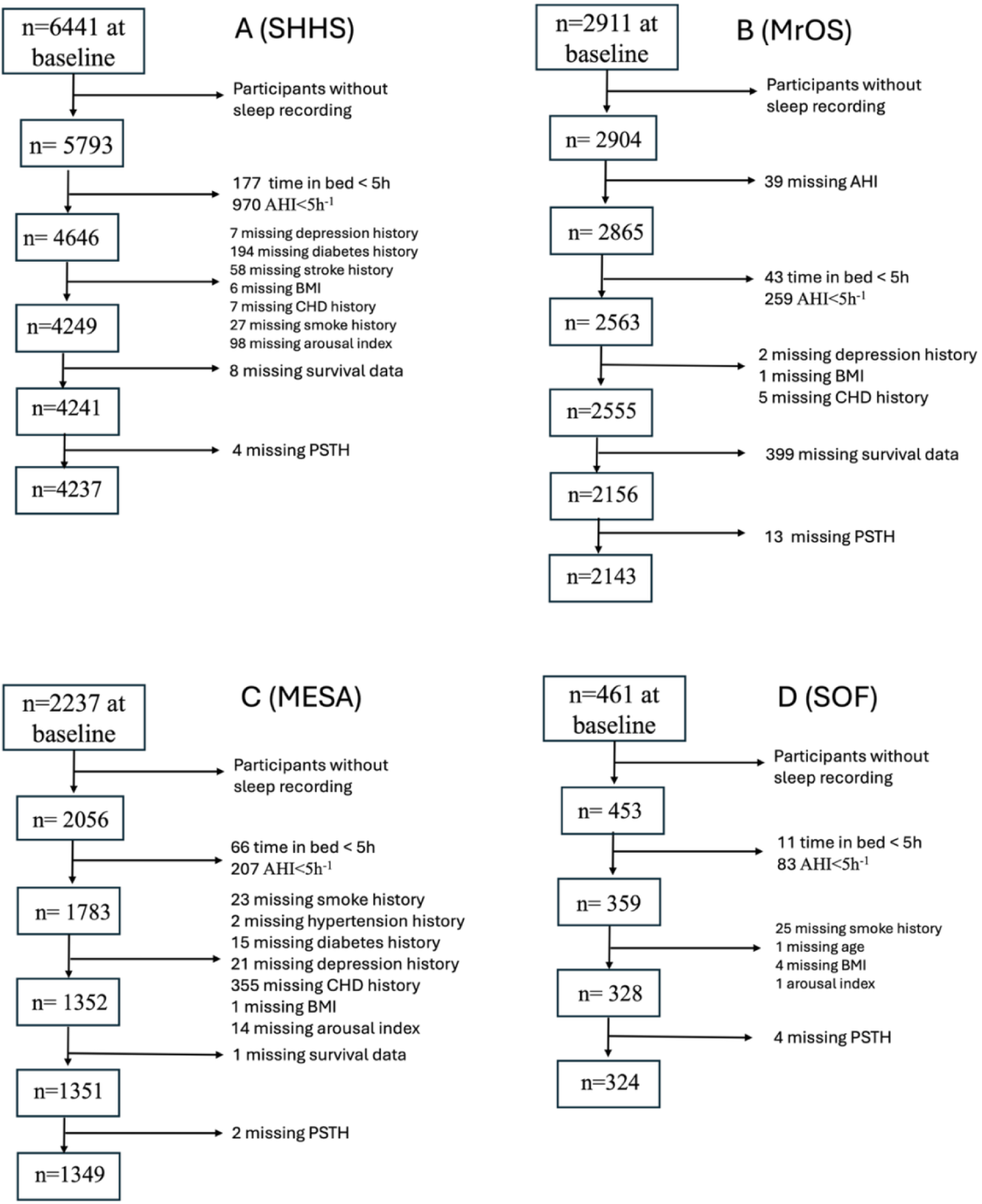
Cohort diagram detailing inclusion and exclusion criteria. The flow diagram detailing inclusion and exclusion criteria for (A) SHHS, (B) MrOS, (C) MESA, (D) SOF. **Alt text:** Flow diagram showing participant selection and exclusions in four cohorts (SHHS, MrOS, MESA, and SOF), from baseline samples to final analytic samples.

### Outcome ascertainment

We investigated two outcomes, all-cause mortality and CVD mortality in each cohort. Note that the outcomes in SHHS were ascertained through its ancillary studies, specifically the Atherosclerosis Risk in Communities (ARIC: 1987-1989) and Framingham Heart Study Offspring I (FHS-OS, i.e., Gen 2: 1995-1998) (21). Details of outcome ascertainment in each cohort are provided in Supplemental method 1 in the supplement. Briefly, all-cause mortality was ascertained through death certificates with or without panel review. CVD mortality was ascertained by panel review.

### Covariates

Covariates were included to address potential confounding, including age at the sleep study, sex, race/ethnicity, BMI, current smoking, mean arterial pressure (MAP), and benzodiazepine use, presence of hypertension, diabetes, depression, coronary heart disease (CHD), and chronic obstructive pulmonary disease (COPD). Age and BMI were modeled using a 3-knot restricted cubic spline at the 5th, 50th, and 95th percentiles. In addition to the primary multivariable models, AHI, arousal index (ArI), and hypoxic burden (HB) were each added separately to assess whether PT predicted mortality independently of these established indices. The details on variable definitions, data sources, and missing rates are provided in the Supplement.

### Post-apnea/hypopnea arousal dynamics

We computed a peri-stimulus time histogram (PSTH) to quantify the time-locked arousal dynamics at the termination of apnea or hypopnea events (Figure 2) for each sleep record. From the normalized PSTH, we extracted three features: peak height (PH), representing the maximal instantaneous probability of arousal; peak time (PT), representing the latency of the peak response relative to apnea termination; and the Area Under Curve (AUC), representing the cumulative probability over the post-event window. To quantify the overlap between these extracted features and standard sleep metrics, we calculated the coefficient of determination (𝑅^2^) for models that included apnea-hypopnea index (AHI), arousal index (ArI), hypoxic burden (HB) (9), total sleep time, sleep efficiency, and arousal burden (AB)(22). We extracted HB and AB using the methods reported in the original study (3,4). We derived a baseline arousal probability 𝑝_baseline_ from the total sleep period, independent of respiratory events (i.e., the overall fraction of 1-second epochs containing any arousal). We normalized each PSTH by subtracting 𝑝_baseline_.

**Figure 2.**
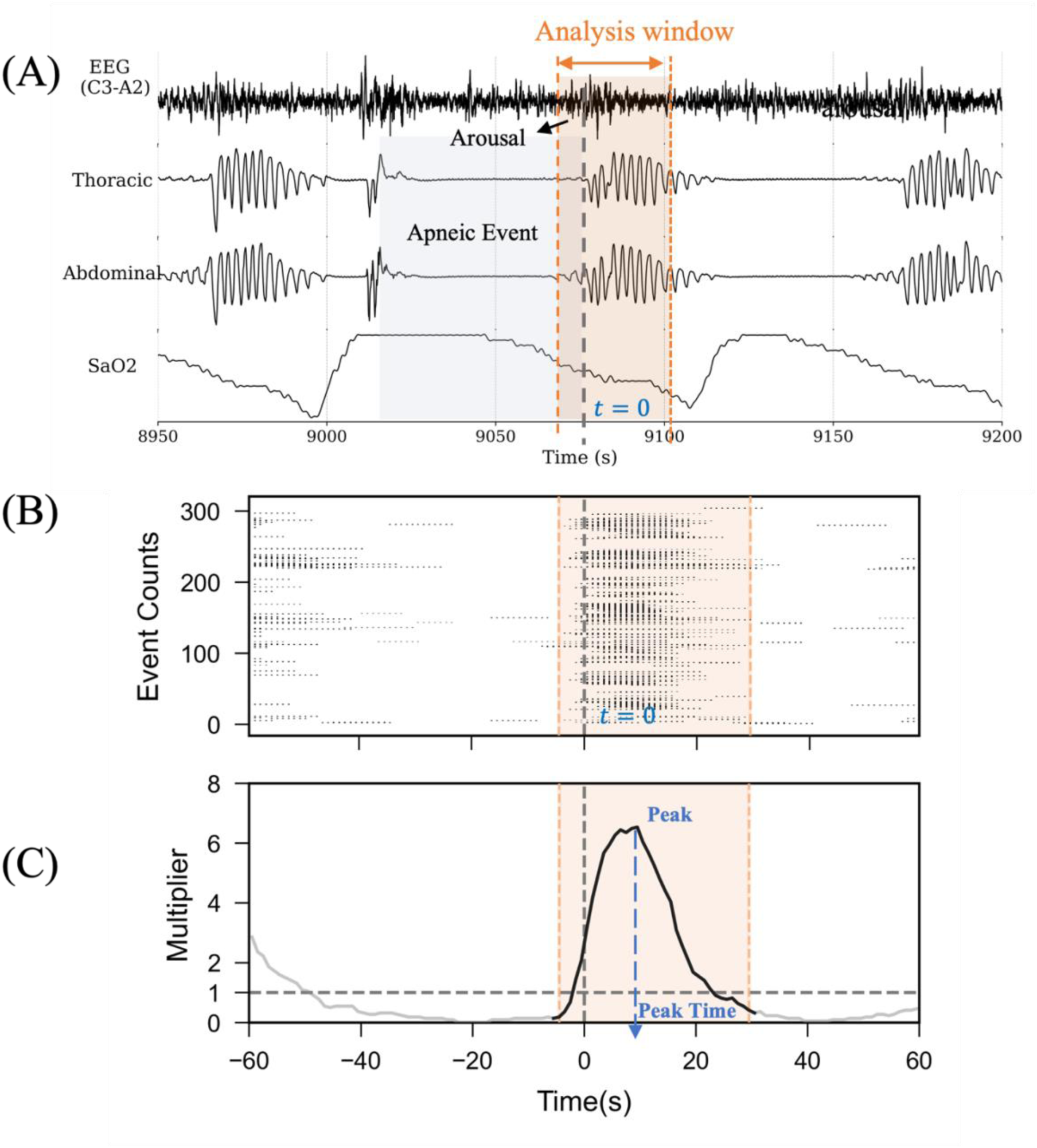
Conceptual framework of the peri-stimuli time histogram (PSTH) and derived peak time (PT). (A) Example of a single apnea event (blue shaded area) ending at time 0, with an arousal followed (orange shaded area), as well as the 35-s window (−5 s to +30 s) (dashed orange lines) used to detect arousals. (B) Distribution of all apneic/hypopnea events for a single participant (82-year-old female), each aligned at event termination (time 0). (C) PSTH curve generated by aggregating arousal occurrences (onset or continuation) within each 1-second bin across all events during the sleep period. Peak height is the highest probability of the arousal reached in the searching window, peak time is the corresponding timing relative to apnea termination, and the area under the curve above baseline is computed as the integral of the curve above baseline (y=1) across the 35-s window. These features characterize an individual’s response to respiratory events. **Alt text:** Multi-panel diagram showing a representative apnea event, alignment of repeated events at event termination, and the resulting peri-stimulus time histogram used to derive peak height, peak time, and area under the curve.

### Cohort-specific analysis

We stratified each PSTH feature into quartiles and compared survival across groups with Kaplan-Meier curves and log-rank tests. We used Cox Proportional Hazards models to assess the association (hazard ratio, HR) between continuous PSTH features and mortality outcomes in each cohort. We fit the model separately on each cohort. We stratified results by sex, given the sex differences in the association between arousal-related biomarker and mortality in prior work (22).

### Individual participant data meta-analysis

Random-effects meta-analysis was used to obtain pooled estimates of the HR for PT (per 1-second) on mortality across all cohorts, separately by sex and outcome. We combined the cohort-specific beta estimates (from the fully adjusted models) using DerSimonian-Laird random-effects and evaluated heterogeneity using the I^2^ statistic.

## Statistical analysis

The linearity assumption for PSTH features as continuous predictors in Cox proportional hazards models was examined using Martingale residuals and restricted cubic splines (Figure S1 in supplement). Robust (Huber-White sandwich)(23) standard errors were used to accommodate minor departures from proportional hazards, thereby yielding valid estimates of the time-weighted average HR if proportionality was not strictly met.

Results are reported as HR with 95% confidence intervals (CIs). No multiple-comparison correction is made because the three features are derived from the common prespecified PSTH. Two-sided p<0.05 was considered significant. All analyses were performed using Python (lifelines package, version 0.3.0) and R (version 4.3.1), between August 2024 and December 2025.

## Results

### Cohort characteristics

A total of 8,053 individuals from four community-based cohorts were analyzed. A total of 2,154 participants (26.6%) died, including 548 deaths attributed to cardiovascular disease. As in Table 1, SHHS (n=4,237, aged 64.4 ± 10.7) and MESA (n=1,349, aged 68.8 ± 9.1) include participants primarily in their mid-to-late 60s, while MrOS (n=2,143, aged 76.5 ± 5.5) and SOF (n=324, aged 83.0 ± 2.9) represent older participants. The proportions of male and female participants were comparable in SHHS (51.8% male) and MESA (48.4% male), whereas MrOS and SOF enrolled only male and female participants, respectively. MESA has greater racial and ethnic diversity (36.5% White, 13.1% Chinese American, 24.9% Hispanic, and 25.5% Black/African American) compared to other cohorts. Hypertension prevalence ranged from about 44.6% (SHHS) to 60.5% (SOF). Other cardiometabolic comorbidities and depression also differ across cohorts. Sleep measures indicated mild-to-moderate apnea severity, with median AHI values ranging from 15.7 to 20.6/hour and median ArI ranging from 18.1 to 22.2/hour.

**Table 1.** Baseline characteristics of participants in all cohorts.

| **Variable** | **SHHS**<br>**(n = 4,237)** | **MESA**<br>**(n = 1,349)** | **MrOS**<br>**(n = 2,143)** | **SOF**<br>**(n = 324)** |
| --- | --- | --- | --- | --- |
| Age, year | 64.4 ± 10.7 | 68.8 ± 9.1 | 76.5 ± 5.5 | 83.0 ± 2.9 |
| Female, n (%) | 2,042 (48.2%) | 696 (51.6%) | 0 (0.0%) | 324 (100.0%) |
| Male, n (%) | 2,195 (51.8%) | 653 (48.4%) | 2,143(100.0%) | 0 (0.0%) |
| Non-White, n (%) | 502 (11.8%) | 857 (63.5%) | 178 (8.3%) | 0 (0.0%) |
| BMI, kg/m<sup>2</sup> | 28.6 ± 5.1 | 28.9 ± 5.4 | 27.2 ± 3.8 | 27.7 ± 4.5 |
| Current Smoking, n (%) | 352 (8.3%) | 89 (6.6%) | 42 (2.0%) | 1 (0.3%) |
| Hypertension, n (%) | 1,889 (44.6%) | 778 (57.7%) | 1,078 (50.3%) | 196 (60.5%) |
| Depression, n (%) | 285 (6.7%) | 188 (13.9%) | 132 (6.2%) | 37 (11.4%) |
| Diabetes, n (%) | 322 (7.6%) | 280 (20.8%) | 294 (13.7%) | 48 (14.8%) |
| Stroke history, n (%) | 147 (3.5%) | - | 80 (3.7%) | 39 (12.0%) |
| CHD history, n (%) | 283 (6.7%) | 26 (1.9%) | 653 (30.5%) | 41 (12.7%) |
| AHI, /hour | 16.2 (9.8-26.6) | 20.6 (12.0-34.8) | 18.7 (11.0-31.4) | 15.7 (9.9-24.5) |
| Arousal index, /hour | 18.1 (13.3-25.1) | 20.4 (14.9-28.8) | 22.2 (16.2-30.0) | 19.2 (14.4-27.9) |
| Hypoxic burden, %minute/hour | 42.1 (26.4-66.7) | 41.6 (24.3-71.5) | 40.6 (25.3-67.6) | 37.1 (23.9-57.0) |
| Peak Time (PT), second | 5.5 (4.5-7.5) | 5.5 (4.5-7.5) | 5.5 (4.5-7.5) | 5.5 (4.5-7.5) |
| Peak Height (PH) | 3.9 (3.2-4.9) | 4.2 (3.4-5.1) | 5.3 (4.2-6.6) | 4.6 (3.7-5.7) |
| Area Under Curve Above Baseline (AUC) | 36.9 (27.5-50.0) | 44.9 (33.0-60.8) | 58.0 (42.8-78.2) | 46.1 (35.0-66.9) |
| All-cause deaths, n (%) | 996 (23.5%) | 206 (15.3%) | 863 (40.3%) | 89 (27.5%) |
| CVD deaths, n (%) | 292 (6.9%) | 58 (4.3%) | 158 (7.4%) | 40 (12.3%) |
| Follow-up, year | 10.9 ± 3.1 | 9.3 ± 1.7 | 8.8 ± 2.8 | 6.4 ± 1.7 |

\*Values are presented as mean ± SD for normally distributed continuous variables or as median (interquartile range) for skewed variables (e.g., Peak Time). Categorical variables are shown as number (%). Abbreviations: SHHS, Sleep Heart Health Study; MESA, Multi-Ethnic Study of Atherosclerosis; MrOS, Osteoporotic Fractures in Men Study; SOF, Study of Osteoporotic Fractures; BMI, body mass index; AHI, apnea-hypopnea index; CHD, coronary heart disease; CVD, cardiovascular disease. SOF: Demographic characteristics are shown for all-cause mortality. For CVD mortality, the sample size is n = 324 after excluding missing cause of death. Because of the relatively small SOF sample, all-cause and CVD mortality are analyzed separately.

Although all cohorts experienced notable nocturnal oxygen desaturation (median HB 37.1-42.1% minute/hour), their median PT remained relatively consistent at around 5.5 seconds.

### Population-averaged PSTH reveals a unimodal pattern

Figure 3 shows the population-averaged PSTH curves for the four cohorts (SHHS, MrOS, SOF, and MESA). Each cohort exhibits a consistent unimodal pattern, with a peak in arousal activity for most participants. The same unimodal pattern holds in all strata (Figure S2-S4 in the Supplement). A small subset of participants (n=32 in SHHS, n=11 in MESA, n=5 in MrOS, and n=6 in SOF) did not display a PSTH peak, as their arousal probabilities within each post-apnea/hypopnea bin never exceeded the full-night arousal probability (all p > 0.05, binomial tests). Rare instances of negative PT indicate that arousal peaked before apnea termination (n = 13 in SHHS, n=5 in MrOS, n=5 in MESA, and n=1 in SOF).

**Figure 3.**
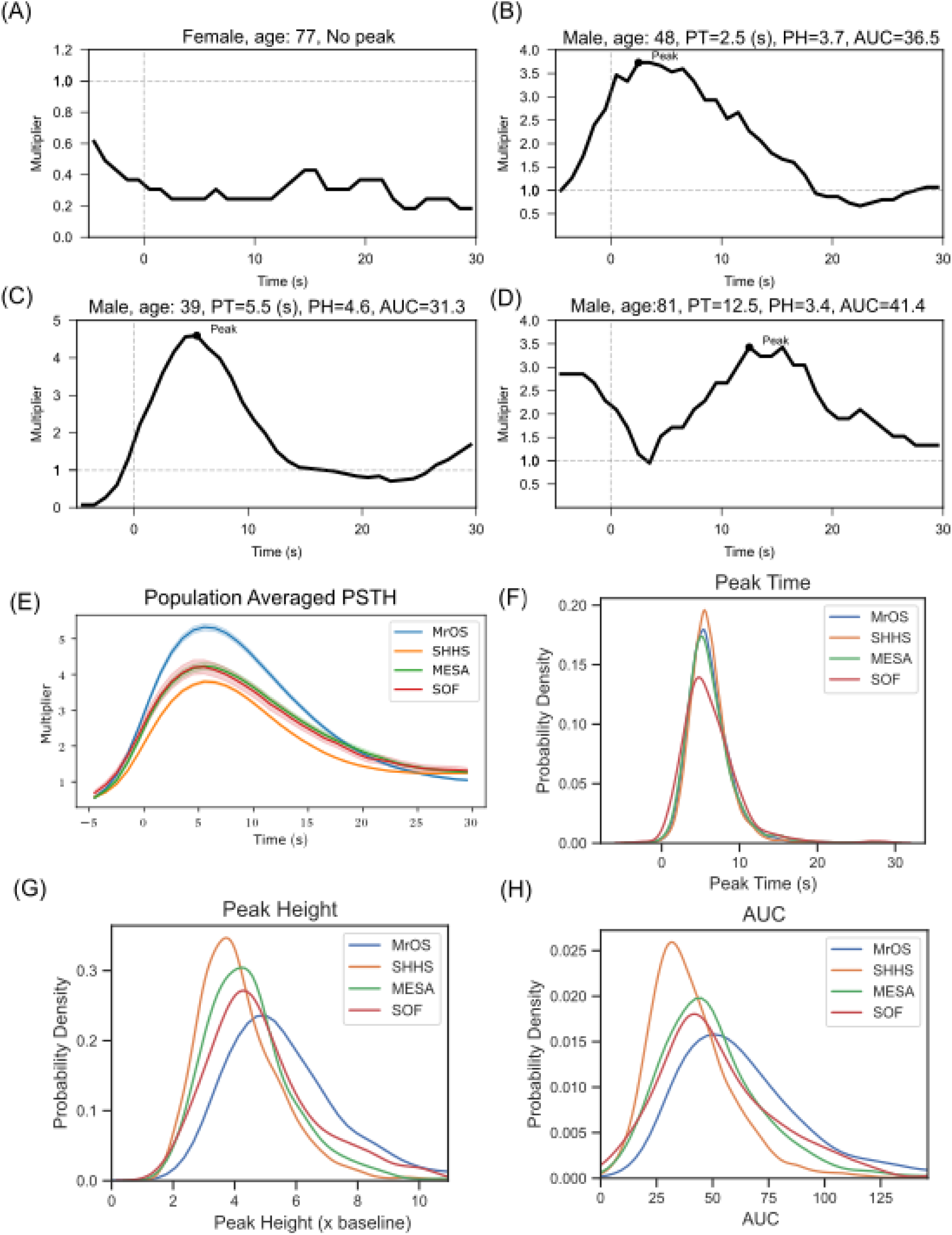
Illustrative examples of individual peri-stimulus time histograms (PSTHs) time-aligned to apnea end and cohort-wide distributions of the PSTH peak. (A) Participant with no definable peak, indicating that no time bin rose significantly above baseline probability. (B) Participant with an early peak time (PT) around 2.5 s (C) Participant exhibiting a mid-range PT (∼5.5 s). (D) Participant with delayed PT (∼12.5 s). The x-axis spans −5 s to +30 s relative to apnea termination. (E) Population-averaged PSTH curves for the four cohorts (MrOS, SHHS, MESA, SOF) show a unimodal pattern in arousal probability. (F) Distribution of PT indicates a right-skewed distribution in all cohorts (Shapiro-Wilk test, p<0.05), with median PT values typically between ∼4.5 s and 5.5 s. (G) Distribution of PH deviated from normality (Shapiro-Wilk test, p<0.05), with a median value from 3.9 to 5.3 times the baseline across cohorts. (H) Distribution of AUC deviated from normality (Shapiro-Wilk test, p<0.05). The median values range from 37.3 to 58.4 across cohorts **Alt text:** Multi-panel figure showing examples of peri-stimulus time histograms with no peak, early peak, intermediate peak, and delayed peak, along with cohort-level average PSTH curves and distributions of peak time, peak height, and area under the curve.

Peak time (PT), which represents the latency of maximal arousal probability relative to apnea termination, was significantly later during rapid-eye-movement (REM) sleep than during non-REM (NREM) sleep in MESA, MrOS, and SHHS (all p<0.001; Cohen’s d = 0.28-0.34). In SOF, this difference was not significant (p=0.28). REM sleep also showed lower PH and AUC than NREM sleep in SHHS and MrOS (all p < 0.001). Effect sizes were small to moderate (Cohen’s d =-0.38 to-0.21 for PH and-0.42 to-0.21 for AUC).

In SHHS, males exhibited slightly higher PH (p<0.001, Cohen’s d=0.13) and AUC (p=0.04, Cohen’s d = 0.04) than females. However, the effect size (i.e., Cohen’s d 0.04 - 0.13) was small, limiting their clinical relevance. No sex differences were detected in the MESA cohort (all p>0.05).

To better contextualize the PT phenotype, we examined demographic and clinical correlates of PT using multivariable linear regression in the SHHS cohort (Table S2). Increasing age was associated with later PT in males (β=0.03 [0.02–0.04] per year; p<0.001) and females (β=0.02 [0.00–0.03]; p=0.028). Higher BMI was also associated with later PT in the overall sample (β=0.04 [0.02–0.06] per kg/m²; p<0.001), as well as in males (β=0.04 [0.02–0.07]; p=0.001) and females (β=0.04 [0.01–0.07]; p=0.005).

Cardiovascular disease history was associated with later PT in males (β=0.41 [0.14–0.68]; p=0.003), but not in females (β=0.39 [−0.05–0.84]; p=0.084). In contrast, higher AHI was associated with a slightly earlier PT (β range, −0.03 to −0.01 per events/hour; all p<0.001). Benzodiazepine use, COPD, and MAP were not significantly associated with PT. Additional sex-specific estimates and corresponding analyses for PH and AUC are provided in Tables S2–S4 in the Online Supplement.

Although several covariates were statistically significant predictors of PT, PH, and AUC, they explained only a modest proportion of overall variability (model R², 0.01–0.06). PT also showed little overlap with PH or AUC (all R² < 0.05) and only weak correlations with conventional metrics of respiratory disturbance, hypoxemia, and sleep fragmentation, including the apnea-hypopnea index, hypoxic burden, arousal index, and arousal burden. In contrast, PH and AUC shared substantial variance (R² = 0.82). Together, these findings suggest that PT captures a physiologic dimension not reflected by conventional sleep apnea severity metrics.

### Arousal peak time after apnea/hypopnea termination is associated with mortality

Peak height (PH) and area under the curve (AUC) were not significantly associated with mortality. Accordingly, Kaplan-Meier analyses for PT are shown in Figures 4 and 5, whereas those for PH and AUC are provided only in Figures S5-S8 in the Online Supplement. Subsequent analyses focused on PT.

**Figure 4.**
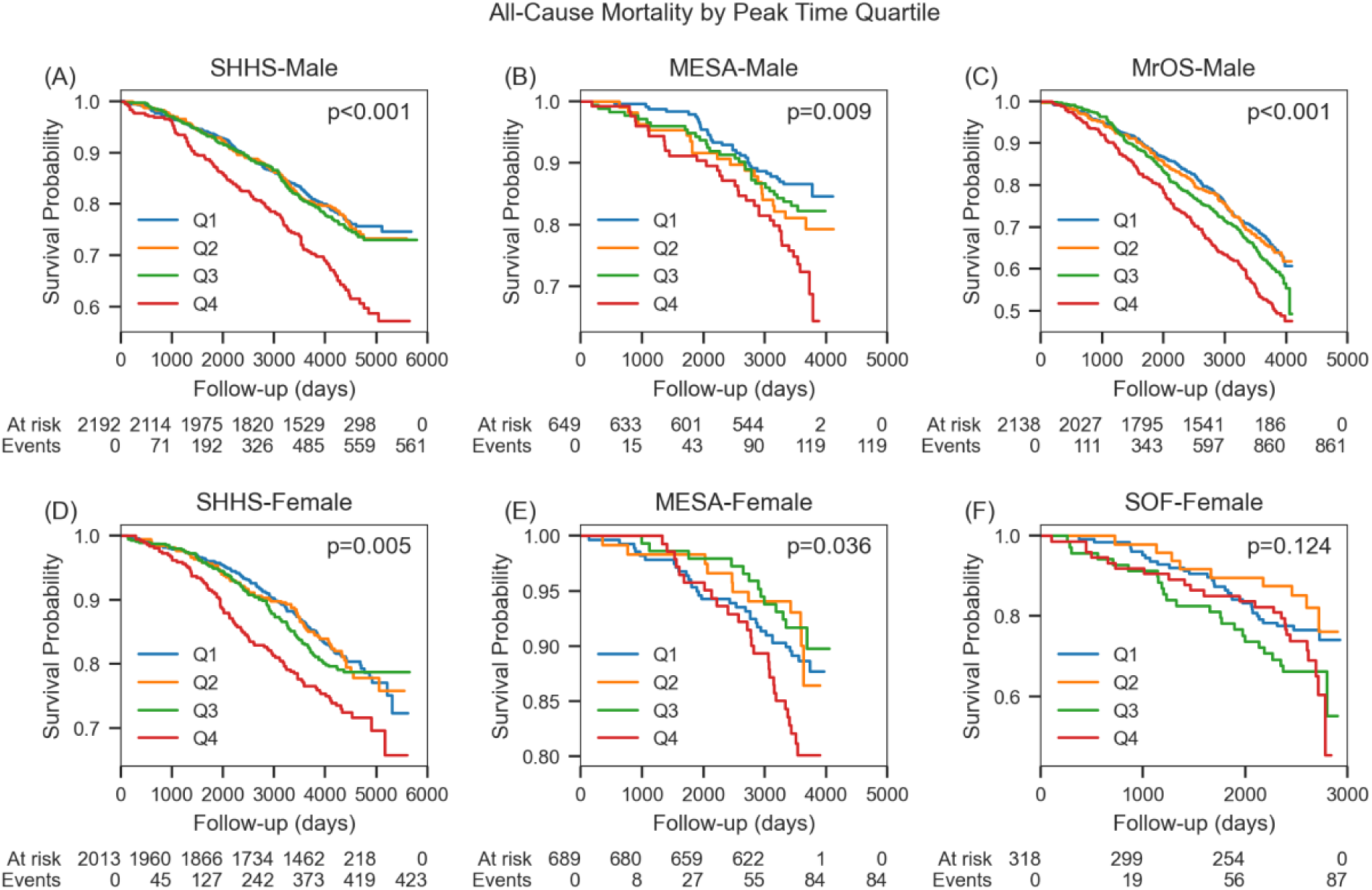
All-Cause Mortality Kaplan-Meier Curves by PT Quartiles. Panels (A)–(C) correspond to males (SHHS, MESA, MrOS), and panels (D)–(F) to females (SHHS, MESA, SOF). The x-axis indicates follow-up time (days), and the y-axis depicts the probability of survival from any cause. Numbers below each panel show participants at risk (“At risk”) and the cumulative number of events (“Events”) at designated intervals. Log-rank tests were performed to compare survival differences among the four PT quartiles, with p-values displayed in the top-right corners of each plot. **Alt text:** Kaplan-Meier plots of all-cause survival stratified by peak time quartiles across six sex-specific cohort panels, with numbers at risk, cumulative events, and log-rank p-values.

**Figure 5.**
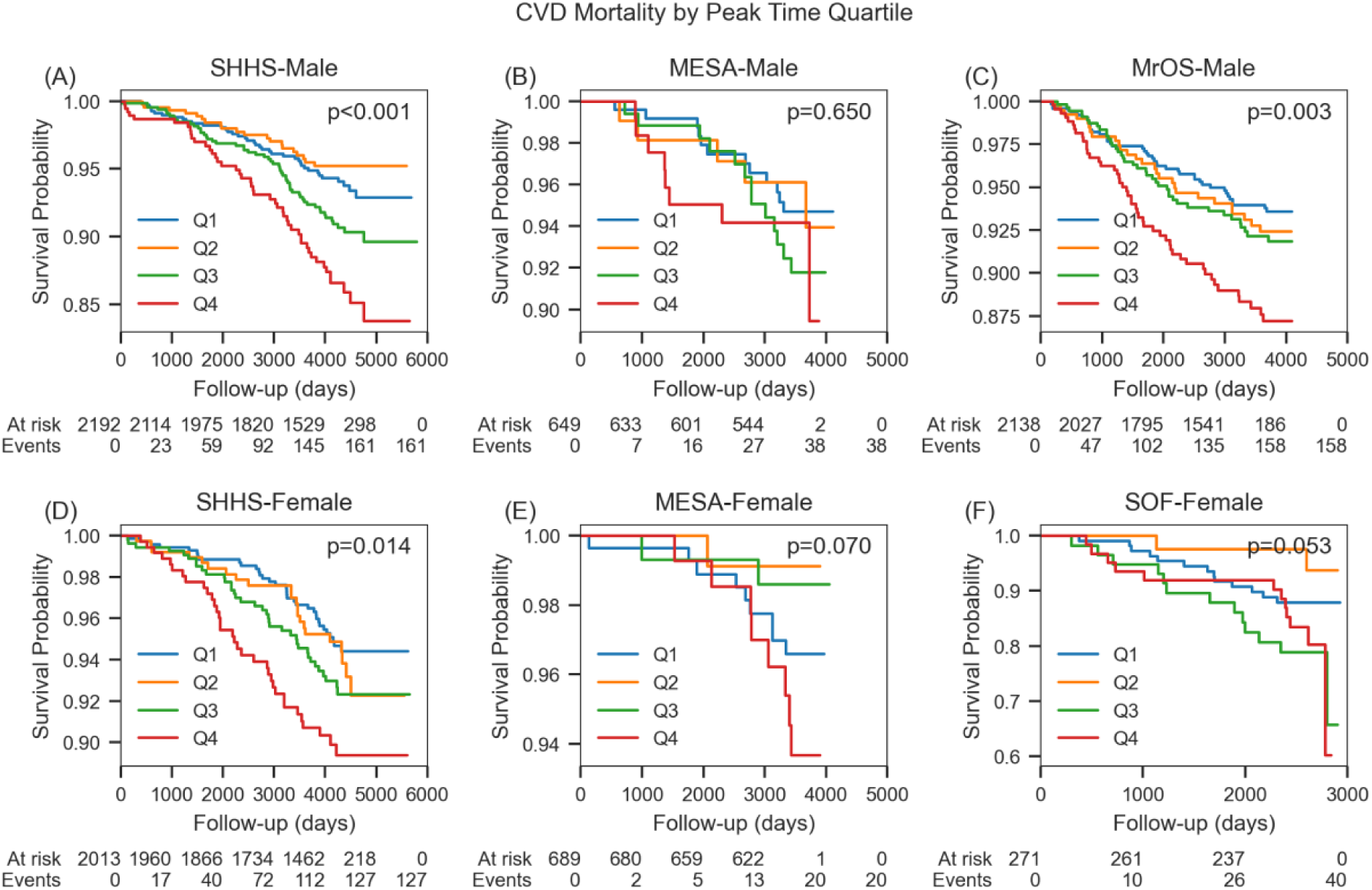
CVD Mortality Kaplan-Meier Curves by PT Quartiles. Panels (A)–(C) correspond to males (SHHS, MESA, MrOS), and panels (D)–(F) to females (SHHS, MESA, SOF). The x-axis indicates follow-up time (days), and the y-axis depicts the CVD event-free survival. Numbers below each panel show participants at risk (“At risk”) and the cumulative number of events (“Events”) at designated intervals. Log-rank tests were performed to compare survival differences among the four PT quartiles, with p-values displayed in the top-right corners of each plot. **Alt text:** Kaplan-Meier plots of cardiovascular event-free survival stratified by peak time quartiles across six sex-specific cohort panels, with numbers at risk, cumulative events, and log-rank p-values.

In Table 2, in SHHS, each 1-second delay in PT was associated with 3% higher all-cause mortality in females (HR=1.03, [1.00-1.06], p=0.023), but non-significant in males (HR=1.03, [1.00-1.06], p=0.09). The association was also evident in MrOS, with an approximately 5% increase in risk per second (HR=1.05 [1.02-1.07], p<0.001) delay in PT. Females in SOF showed a significant association between PT and all-cause mortality (HR=1.07, [1.01 - 1.13], p=0.029). For CVD mortality, both males and females in SHHS showed moderate but significant associations between delayed PT and higher CVD death risk (males: HR=1.06 [1.01-1.11], p=0.031; females:1.06 [1.01-1.11], p=0.014). In MrOS, delayed PT was associated with increased CVD mortality risk (HR=1.06 [1.01-1.11], p=0.028). In contrast, MESA did not show an association between PT and all-cause or CVD mortality (p>0.05). Importantly, the significant associations observed in SHHS, MrOS, and SOF persisted, although slightly attenuated, after additional adjustment for key sleep parameters including AHI, ArI, and HB.

**Table 2.** PT as a continuous value vs. mortality.

|  |  | **All-cause mortality** |  | **CVD-mortality** |  |
| --- | --- | --- | --- | --- | --- |
|  | **Models** | **HR (95%CI)** | **P-value** | **HR (95%CI)** | **P-value** |
| SHHS Male | PT | 1.03 (1.00-1.06) | 0.090 | **1.06 (1.01-1.11)** | **0.031** |
|  | PT+Arl | 1.03 (1.00-1.06) | 0.077 | **1.05 (1.00-1.11)** | **0.042** |
|  | PT+AHI | 1.03 (1.00-1.06) | 0.063 | **1.06 (1.01-1.11)** | **0.030** |
|  | PT+HB | 1.03 (1.00-1.06) | 0.078 | **1.06 (1.01-1.11)** | **0.028** |
| SHHS Female | PT | **1.03 (1.00-1.06)** | **0.024** | **1.06 (1.01-1.11)** | **0.014** |
|  | PT+Arl | **1.03 (1.00-1.06)** | **0.023** | **1.06 (1.01-1.11)** | **0.013** |
|  | PT+AHI | **1.03 (1.00-1.06)** | **0.023** | **1.06 (1.01-1.11)** | **0.014** |
|  | PT+HB | **1.03 (1.00-1.06)** | **0.023** | **1.06 (1.01-1.11)** | **0.014** |
| MESA Male | PT | 1.04 (0.98-1.10) | 0.159 | 1.00 (0.91-1.10) | 0.977 |
|  | PT+Arl | 1.04 (0.98-1.11) | 0.188 | 1.00 (0.91-1.10) | 0.940 |
|  | PT+AHI | 1.05 (0.99-1.11) | 0.116 | 1.01 (0.91-1.12) | 0.854 |
|  | PT+HB | 1.04 (0.99-1.11) | 0.142 | 1.01 (0.91-1.12) | 0.845 |
| MESA Female | PT | 0.99 (0.93-1.06) | 0.816 | 0.94 (0.82-1.08) | 0.374 |
|  | PT+Arl | 1.00 (0.93-1.07) | 0.961 | 0.95 (0.82-1.10) | 0.483 |
|  | PT+AHI | 0.99 (0.93-1.07) | 0.879 | 0.94 (0.80-1.10) | 0.409 |
|  | PT+HB | 1.00 (0.93-1.07) | 0.910 | 0.94 (0.82-1.08) | 0.372 |
| SOF Female | PT | **1.07 (1.01-1.13)** | **0.029** | 1.07 (0.97-1.18) | 0.205 |
|  | PT+Arl | **1.08 (1.02-1.14)** | **0.009** | 1.10 (1.00-1.21) | 0.061 |
|  | PT+AHI | **1.08 (1.02-1.14)** | **0.010** | 1.09 (0.98-1.23) | 0.119 |
|  | PT+HB | **1.07 (1.02-1.14)** | **0.013** | 1.08 (0.97-1.21) | 0.168 |
| MrOS Male | PT | **1.05 (1.02-1.07)** | **<0.001** | **1.06 (1.01-1.11)** | **0.028** |
|  | PT+Arl | **1.05 (1.02-1.07)** | **<0.001** | **1.06 (1.01-1.11)** | **0.024** |
|  | PT+AHI | **1.05 (1.02-1.07)** | **<0.001** | **1.06 (1.01-1.11)** | **0.027** |
|  | PT+HB | **1.05 (1.02-1.07)** | **<0.001** | **1.06 (1.01-1.11)** | **0.026** |

\*Values represent the hazard ratio per 1-second increase in Peak Time (PT). Bolded values indicate statistically significant associations based on the Wald test. AHI: Apnea-Hypopnea Index; ArI: Arousal Index; HB: Hypoxic Burden.

Figure 6 presents the meta-analysis results across subgroups. In males (SHHS, MESA, and MrOS), each 1-second delay in PT was associated with a 4% higher risk of all-cause mortality (pooled HR=1.04, [1.02-1.06], p<0.001) and a 5% higher risk of CVD mortality (pooled HR=1.05 [1.02-1.08], p=0.004) with no evidence of heterogeneity (I^2^=0.0% for both outcomes). Given the 3-second PT IQR in Table 1, a 3-second delay in PT was associated with a 12% higher risk of all-cause mortality and a 15% higher risk of CVD mortality. In females (SHHS, MESA, and SOF), each 1-second delay in PT was associated with a 3% higher risk of all-cause mortality and a higher cohort heterogeneity (pooled HR =1.03 [1.00–1.06], p=0.04, I^2^=23.4%), while not associated with CVD mortality (pooled HR=1.04 [0.99-1.10], p=0.13, I^2^=25.3%). A 3-second delay in PT was associated with a 9% higher risk of all-cause mortality.

**Figure 6.**
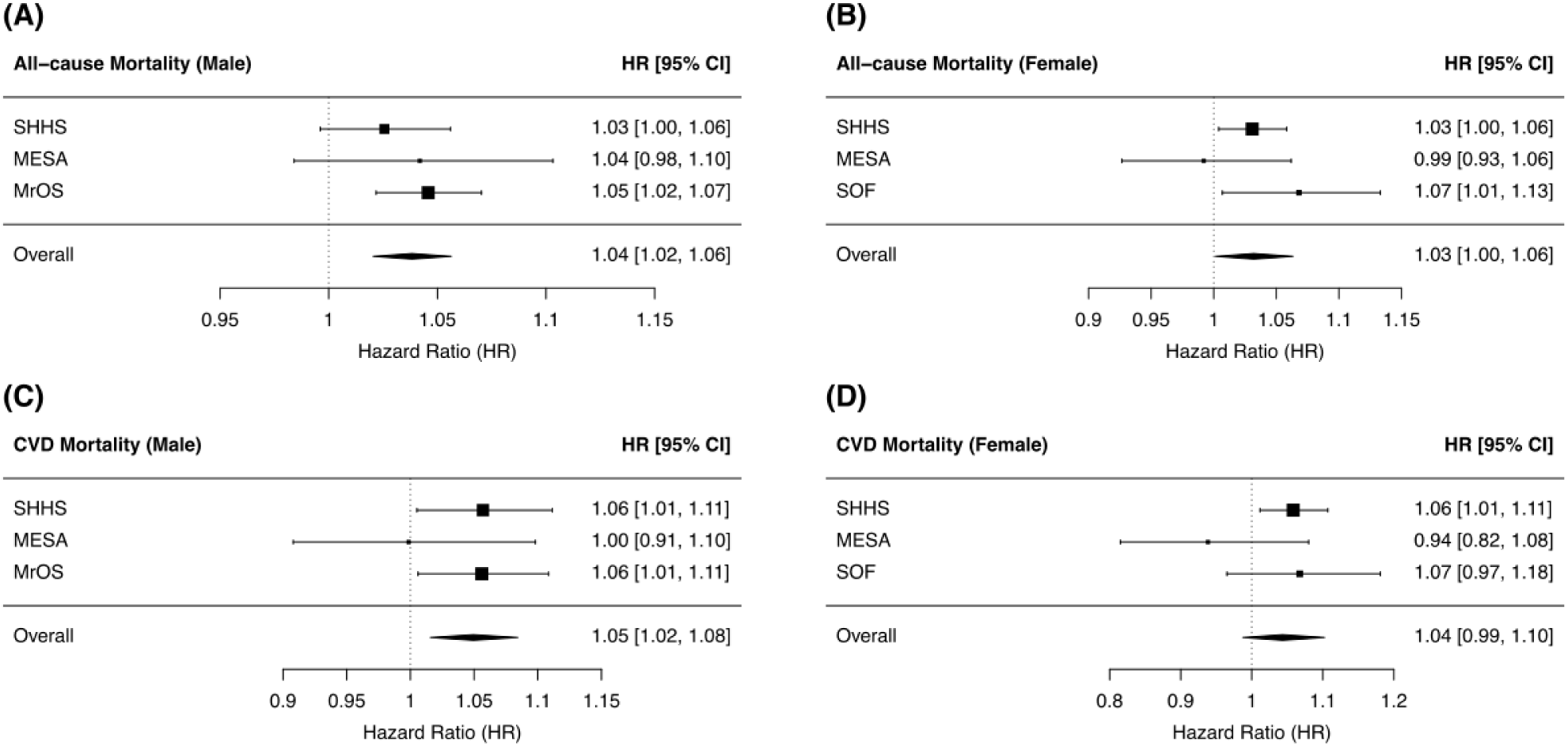
Meta-analyses of the association between peak time (PT) and mortality risk. Forest plots show pooled hazard ratios (HRs) and 95% confidence intervals (CIs) for the association between PT and (A) all-cause mortality in males, (B) all-cause mortality in females, (C) cardiovascular (CVD) mortality in males, and (D) CVD mortality in females. Each HR represents a cohort-specific estimate derived from multivariable-adjusted Cox Proportional Hazards models. Pooled estimates were calculated using random-effects meta-analysis with the DerSimonian–Laird (DL) estimator. No significant heterogeneity was observed in male subgroups (I² = 0.0%), while the female subgroup showed moderate heterogeneity (all-cause mortality: I² = 23.4%; CVD mortality: I² = 25.3%). **Alt text:** Forest plots showing cohort-specific and pooled hazard ratios for the association between delayed peak time and all-cause or cardiovascular mortality in males and females.

To assess potential nonlinearity, we additionally categorized PT into quartiles (Table S8). In SHHS females, the highest quartile (Q4) was associated with higher all-cause mortality (HR=1.32 [95% CI, 1.02–1.71]; p=0.036) and cardiovascular mortality (HR=1.75 [1.08–2.83]; p=0.023) relative to Q1. Similarly, in MrOS, Q4 was associated with higher all-cause mortality (HR=1.31 [1.09–1.57]; p=0.004) and cardiovascular mortality (HR=1.69 [1.09–2.60]; p=0.018). Corresponding quartile-based associations were not statistically significant in MESA or SOF. Model-adjusted survival curves by PT quartiles are shown in Figures S13 and S14 in the Online Supplement.

### Associations with mortality were observed for NREM-PT but not REM-PT

Table S1 presents the results of PT separately derived from apneas and hypopneas during REM and NREM sleep. PT-REM was near null across all subgroups (all p >0.05). PT-NREM showed significant associations in SHHS females (all-cause mortality: HR=1.03 [1.01–1.06], p=0.006; CVD mortality: HR=1.06 [1.02–1.11], p=0.006) and in MrOS males for all-cause mortality (HR=1.04 [1.02–1.06], p<0.001), but not for CVD mortality in MrOS (HR=1.02 [0.97–1.07], p=0.41). SHHS males showed no associations for PT-NREM, and SOF females showed no significant associations.

## Discussion

We used the stressor-response framework (11) to characterize apnea-arousal dynamics using the peri-stimulus time histogram (PSTH) aligned to apnea/hypopnea event termination, yielding three metrics: Peak Time (PT), Peak Height (PH), and Area Under the Curve (AUC). PT (the latency of highest arousal probability relative to apnea/hypopnea termination) emerged as a predictor of health outcomes. In contrast, PH (the maximal instantaneous probability) and AUC (cumulative probability) were not significantly associated with mortality. In pooled meta-analysis across four datasets, longer PT (i.e., delayed arousal) was associated with higher all-cause and cardiovascular mortality and with higher CVD mortality in males, but not in females.

Cohort-specific estimates were heterogeneous but were generally directionally concordant in SHHS, SOF, and MrOS, with largely null associations in MESA. Sex-specific patterns were also observed. Heterogeneity was lower in males than in females (I²=0% vs ∼23–25%). The results held after additional adjustment for AHI, ArI, and HB. Our results suggest that the timing of arousal may be as important as its frequency.

Arousals are coupled to the termination of apneic events (24). A large-scale analysis (25) quantified the typical distribution of arousals around apnea/hypopnea events. In over 2 million apneic events and ∼1.6 million arousals, ∼90% of apnea-related arousal occurred within 4 seconds before to 9 seconds after the apnea end, with the peak probability of arousal onset essentially coinciding with apnea end, while arousal duration midpoint probability peaked at 6 seconds after apnea end. However, the results in (25) also showed that a non-trivial minority of apneas were followed by delayed arousal up to 10-15 seconds later. For example, high loop-gain-related arousals crest the arousal complex and occur later than airway-opening related arousals that are related to pure obstructive events. Because high loop gain involves carotid body chemoreflex hypersensitivity, there could be greater sympathetic discharge. Within the stressor-response framework, it is reasonable to hypothesize that a later PT could coincide with larger post-event sympathetic surges and blood-pressure spikes that are detrimental to the vascular endothelium (24, 26). In our study, the PT metric captures this individual-specific variability in the temporal relationship between arousal and apnea.

In our study, the apnea-end-locked arousal peak time was not significantly associated with HB. Therefore, the observed association between a longer PT and mortality does not appear to be related to greater hypoxemia. PT is a temporal measure referenced to the end of the respiratory event and is independent of the duration of the obstructive event as incorporated in HB.

The temporal relationship between airway reopening and arousal varies between sleep stages (26). A notable finding from our study is that the predictive value of PT was specific to NREM sleep. In our analyses, we found that PT-NREM is significantly shorter than PT-REM within an individual. Moreover, delayed arousal timing during NREM (longer PT-NREM) was consistently associated with higher mortality risk, whereas PT-REM was not significantly associated with outcomes. This divergence likely reflects fundamental physiological differences between NREM and REM sleep in the context of OSA. During NREM, especially in the deeper stages (N3), the brain is less responsive and has higher arousal thresholds. As a result, obstructive events in NREM can persist for an extended period until a substantial arousal eventually terminates the apnea. In individuals with an inherently high arousal threshold, NREM apneas may become long and severe before arousal occurs, leading to high PT-NREM and greater cardiovascular stress. In REM sleep, however, the situation is more complex. REM sleep often shows greater respiratory variability and reduced muscle tone (atonia). In healthy subjects, REM can display a comparable or even slightly lower arousal threshold than lighter NREM stages. However, in OSA patients, REM-related hypotonia may amplify airway collapse, prolong events, and raise the effective threshold required for arousal. This might partially explain the longer PT-REM compared to PT-NREM. Meanwhile, REM is less chemosensitive than NREM, leading to apneas mainly of obstructive origin.

Importantly, several factors may make PT-REM less physiologically relevant. First, the PSTH may underrepresent the actual occurrence of arousal in REM, as some REM apneas may resolve without triggering a cortical event that meets the EEG arousal standard (per scoring rules requiring both EEG and chin EMG changes). Second, REM sleep constitutes a smaller fraction of total sleep (15-25%) (27), thereby reducing the statistical power for REM-specific PT analyses. In essence, PT-NREM likely captures individual differences in arousal threshold, whereas PT-REM may be sensitive to other factors.

Our findings also align with a broader shift in sleep research from count-based measures toward multi-dimensional, comprehensive markers. Recent large cohort studies have shown that metrics that account for more composite information, such as the depth of hypoxemia and the duration of arousal, have superior predictive value than the AHI. For instance, hypoxic burden, which captures the total depth and duration of oxygen desaturation during all apneas, has emerged as an important indicator of CVD risk (9). Arousal burden, which measures the total time spent in arousal during sleep, was a predictor of long-term mortality in over 8,000 older adults (22). Likewise, the apnea-induced heart rate response (10) is important. The magnitude of the pulse rate surge provoked by each respiratory event independently predicts future cardiovascular events and mortality. Collectively, these prior studies show a trend from characterizing OSA severity solely in a global view to nuanced information at the event level. PT complements the above biomarkers by quantifying the temporal relationship between arousal and apnea. These biomarkers may paint a more complete picture of OSA characterization than AHI alone.

### Strengths and Limitations

This study has several strengths: 1) it included four community-based cohorts with in-home polysomnography and longitudinal follow-up; 2) the individual participant data meta-analysis enabled cohort-specific and pooled estimates; and 3) the association of delayed arousal peak timing with mortality remained after adjustment for apnea-hypopnea index, arousal index, and hypoxic burden. However, the study also had several limitations. First, its observational design does not permit causal inference. A longer peak time may reflect multiple unmeasured processes, including a higher arousal threshold, delayed airway reopening, impaired chemosensitivity, or delayed neural conduction. Second, arousal timing may differ by respiratory event type, including obstructive events, central events, and Cheyne-Stokes respiration. Given the clinical links between central events/Cheyne-Stokes respiration and cardiovascular disease, future studies should pre-specify either exclusion of such events or stratified analyses with adjustment for them. Third, the cohorts were composed predominantly of middle-aged and older adults, which may limit generalizability; younger or more diverse populations may show different peak time-risk relationships. Finally, peak time depends on human scoring of sleep stage, respiratory events, and electroencephalographic arousals and may therefore be subject to measurement variability. Automated detection methods may improve precision in future work. Repeat recordings were separated by several years rather than consecutive nights, so some variability is expected. Multi-night studies are therefore needed to better define the short-term test-retest reliability of peak time.

## Future Directions

Additional studies are needed to prospectively validate these findings. Nonetheless, these results suggest that delayed post-apnea/hypopnea arousal timing may help identify a high-risk subgroup of patients with obstructive sleep apnea beyond conventional severity measures. Future work should also determine whether peak time can be measured more reliably with automated methods, whether its prognostic value differs by respiratory event type, and whether it is modifiable with treatment

## Conclusion

This study provides evidence that delayed post-apnea/hypopnea arousal peak timing is associated with increased mortality risk in obstructive sleep apnea. These associations were independent of apnea frequency, arousal burden, and hypoxic burden, and were most evident for all-cause mortality and for cardiovascular mortality in males.

## Data Availability

The individual-level source data underlying this study are not publicly redistributed by the authors because they are subject to cohort-specific governance and data-use restrictions. Qualified investigators may request access through the original cohort data-access mechanisms, subject to approval. Sleep Heart Health Study data are available through the National Sleep Research Resource. Multi-Ethnic Study of Atherosclerosis data are available through BioLINCC. Osteoporotic Fractures in Men Study and Study of Osteoporotic Fractures data are available through MrOS/SOF Online.

https://sleepdata.org/datasets/shhs

https://biolincc.nhlbi.nih.gov/studies/mesa/

https://sofonline.ucsf.edu/

https://mrosonline.ucsf.edu/

## Supplemental method 1. Study Cohorts

The Sleep Heart Health Study (SHHS) is a prospective, community-based cohort study designed to investigate the associations between sleep-disordered breathing (SDB) and cardiovascular and neurocognitive outcomes. Between 1995 and 1998, SHHS recruited 6,441 middle-aged and older adults from existing epidemiological cohorts at 11 sites across the United States, forming the baseline cohort (SHHS 1). Baseline assessments included in-home polysomnography (PSG) and standardized health questionnaires. The primary endpoint evaluated was all-cause mortality. Mortality status was ascertained using multiple concurrent methods, including periodic follow-up interviews, annual questionnaires or telephone contact with participants or next of kin, surveillance of local hospital records and community obituaries, and linkage to the Social Security Administration Death Master File. By April 1, 2006, 1,047 deaths had been confirmed. Cardiovascular disease (CVD) outcomes, including hospitalized acute myocardial infarction (MI), coronary revascularization procedures (e.g., angioplasty, coronary artery bypass surgery), congestive heart failure, coronary heart disease-related death, and angina pectoris, were ascertained through standardized adjudication processes. For SHHS participants recruited from parent cohorts such as ARIC (Atherosclerosis Risk in Communities), CHS (Cardiovascular Health Study), FHS (Framingham Heart Study), and SHS (Strong Heart Study), existing established protocols within these cohorts were utilized to determine CVD outcomes. Participants from Tucson and New York, where parent cohorts lacked established CVD outcome ascertainment, underwent independent ascertainment procedures closely modeled after those used by CHS.

The Osteoporotic Fractures in Men Study (MrOS) is a prospective, community-based cohort study of older males (65 years and above) across multiple clinical centers to examine associations between sleep disorders and health outcomes, including fractures and mortality. Approximately 2,874 participants underwent in-home PSG, with repeated measurements (Visits 1 and 2) enabling assessments of changes in sleep metrics over time. Participants were followed up through 2016 (end of Visit 4). Reported deaths through 2018, including the causes, were confirmed by a centralized review of death certificates. We used the death status at the end of Visit 4, the last visit at which cognitive status was obtained. To ascertain death, participants were contacted every 4 months via postcard to determine vital status. Next of kin were contacted in cases of nonresponse. The MrOS dataset is available on request from MrOS Online (https://mrosonline.ucsf.edu).

The Multi-Ethnic Study of Atherosclerosis (MESA) is a prospective, community-based cohort study of middle-aged and older adults investigating factors associated with the development of subclinical cardiovascular disease and the progression from subclinical to clinical cardiovascular disease. Between 2010 and 2013, 2,237 participants were also enrolled in a Sleep Exam (MESA Sleep), which included full overnight unattended polysomnography, 7-day wrist-worn actigraphy, and a sleep questionnaire. The objectives of the sleep study are to understand how variations in sleep and sleep disorders differ across gender and ethnic groups and how they relate to measures of subclinical atherosclerosis. The participants were followed up on until 2018. All-cause mortality was ascertained using death certificates from hospitals. The cause of death, including CVD, was classified by two physicians independently.

The Study of Osteoporotic Fractures (SOF) is a prospective, community-based cohort study of older females investigating risk factors for fractures. A subset undergoing PSG at Visit 8 to measure sleep-disordered breathing. Visit 9 provided follow-up data. All-cause mortality was ascertained by contacts every 4 months and confirmed with death certificates. Four clinics individually collect SOF mortality data, and a State Registered Certificate of Death is also submitted to the Coordinating Center. Study physicians adjudicated the underlying causes of death, including CVD. The SOF dataset is available on request via SOF Online (https://mrosonline.ucsf.edu). All participants provided written informed consent in each cohort, and each cohort’s protocol was approved by the institutional review board at its respective study site.

## Supplemental method 2. Sleep Staging, Apnea, and Arousal Scoring

For SHHS, MrOS, and SOF, sleep staging was based on the Rechtschaffen & Kales guidelines (1), in which S3 and S4 were combined into N3 to match the newer guidelines.

Apneas were scored if thermistor-based airflow was absent for at least 10 seconds. Hypopneas were identified when there was at least a 30% reduction in airflow (by thermistor or nasal pressure) or thoracoabdominal movement for at least 10 seconds. Events were also linked to associated desaturation to calculate AHI, defined as the number of all apneas plus hypopnea associated with ≥ 3% desaturation or arousal per hour of sleep. For the event-level analysis, we did not exclude tagged hypopnea events without clearly linked ≥3% desaturation.

Arousal was scored as an abrupt shift to a higher EEG frequency lasting at least 3 seconds and starting after at least 10 continuous seconds of sleep. During REM arousals, the EEG frequency shift was accompanied by a simultaneous increase in chin EMG tone lasting over 1 second.

To verify the accuracy of apnea/hypopnea and arousal annotations, we computed EEG spectrograms time-locked to event termination (0 s). We used the C4–A1 for MESA and C3–A2 derivations for other cohorts. Signals were resampled to 128 Hz after anti-aliasing Finite Impulse Response low-pass filtering (Kaiser window), then band-pass filtered (0.3–45 Hz) with a zero-phase elliptic IIR filter (order 16; passband ripple ≤1 dB; stopband attenuation ≥40 dB). For each event, we extracted segments from 5 s before to 30 s after termination and estimated power with Welch’s method (2-s Hann window; 0.5-s step; 75% overlap). Each frequency bin was normalized to its value at 0 s. As a quality check, high-frequency power (>12 Hz) increased around 0 s, consistent with the annotated arousals (see Figure S9). The histogram of apnea/hypopnea event duration across all participants and cohorts is presented in Figure S10 and Figure S11.

## Supplemental method 3. Characterize Post-Apnea Arousal Dynamics Using PSTH

We computed a peri-stimulus time histogram (PSTH) to quantify the arousal dynamics time-locked to the end of apnea or hypopnea events (Figure 2 in the main text). We did not distinguish between obstructive, central, or mixed apneas/hypopneas. For each apnea or hypopnea, we aligned the event at its termination (time 0) and considered a 35-second window (−5 to +30 seconds) divided into 1-second bins. Let 𝑁_𝑒𝑣𝑒𝑛𝑡𝑠_be the total number of apnea/hypopneas for a given participant, and 𝑝_𝑘_ for bin 𝑘 as:

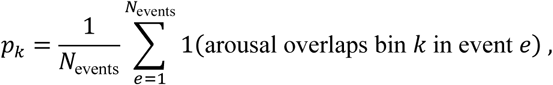

where 𝟏{⋅} is an indicator function that equals 1 if an arousal is present in the bin 𝑘 of event 𝑒. The resulting sequence

{𝑝_𝑘_}_𝑘=1,…,35_is the individual-specific PSTH. We derived a baseline arousal probability 𝑝_baseline_from the total sleep period, independent of respiratory events (i.e., the overall fraction of 1-second epochs containing any arousal). We normalized each PSTH by subtracting 𝑝_baseline_.

From the normalized PSTH, we extracted three features: peak height (PH), peak time (PT), and the Area Under Curve (AUC). PH was defined as the maximum probability 𝑝_𝑘_ that exceeded 𝑝_baseline_ at a statistically significant level (one-sided binomial test multiple-comparison correction using the Benjamini–Hochberg false discovery rate [FDR; q = 0.05], p< 0.05). Peak time was the time when bin 𝑘 corresponding to PH. AUC was computed as the integral of the probability curve above 𝑝_baseline_ across the 35-s window. If no bin exceeded 𝑝_baseline_ at p < 0.05, the participant was assigned no definable PH or PT and was excluded from further analyses. We labelled each 1-s bin by its midpoint to more precisely indicate its span. For example, PT = 3.5 s indicates that the highest arousal probability occurred in the 3–4 s bin after apnea termination. In contrast, PH is expressed as a ratio relative to 𝑝_baseline_.

## Supplemental method 4. Test-Retest Reliability

Test-retest reliability was examined in participants who underwent repeated polysomnography (SHHS and MrOS), focusing on the PSTH shape and its features. Intra-subject versus inter-subject correlations (Pearson’s R) were used to assess consistency of the PSTH waveform. Intraclass correlation coefficient (ICC) was used to quantify the stability of PT, PH, and AUC within individuals across visits.

## Supplemental results 1. Association Between Covariates and Post-Apnea/Hypopnea Arousal Dynamics

Table S3 summarizes the associations between PH and covariates using multivariable linear regression. PH (per times to the baseline) was lower in females than males (β=−0.12 [−0.20, −0.03], p=0.010). Higher BMI was associated with lower PH (β=−0.02 [−0.03, −0.01], p<0.001), and higher AHI was also associated with slightly reduced PH (β=−0.004 [−0.01, −0.00], p=0.006). In females, older age was associated with higher PH (β = 0.01 [0.00, 0.02], p=0.001).

For AUC, older age (β = 0.10, [0.04, 0.15], p=.001) was associated with a higher AUC (Table S4). Being female (β =-3.01, [-4.21,-1.81], p<.001), having a higher BMI (β =-0.22, [-0.34,-0.10], p<.001), and a higher AHI (β =-0.27, [-0.31,-0.23], p<.001) were associated with a lower AUC. In females, significant associations persisted for age (β = 0.15, [0.06, 0.24], p<0.001), BMI (β =-0.25, [-0.41,-0.08], p=.003), and AHI (β =-0.26, [-0.33,-0.19], p<.001). In males, both higher BMI and higher AHI were associated with lower AUC (BMI p=0.048; AHI p<0.001).

In addition, Table S5-7 showed that there is no direct or consistent association between overall or sleep-stage-specific (NREM and REM) PT and excessive daytime sleepiness (ESS) across the study cohorts

## Supplemental results 2. PSTHs Showed Night-To-Night Consistency for Individuals

Two cohorts (SHHS and MrOS) included participants (n=1,988 in SHHS and n=766 in MrOS) with repeated visits separated by several years (SHHS: mean interval = 5.2 ± 0.25 years; MrOS: mean interval 6.5 ± 0.7 years). We evaluated intra-and inter-subject correlation coefficients for PSTH curves across two visits in SHHS and MrOS. As illustrated in Figure S15, PSTHs showed significantly higher intra-subject correlation than inter-subject correlation in both SHHS (intra-subject r = 0.74 vs. inter-subject r = 0.63, p<0.001) and MrOS (intra-subject r = 0.76 vs. inter-subject r = 0.65, p<0.001), indicating that an individual’s PSTH waveform shared relatively consistent patterns despite night-to-night variability. However, the reproducibility of the extracted features was poor. In SHHS, the intraclass correlation coefficient (ICC) was 0.14 [0.10–0.19] for PT, 0.24 [0.18–0.30] for PH, and 0.22 [0.15–0.29] for AUC. The results were similar in MrOS, with corresponding ICCs of 0.14 [0.06–0.22] for PT, 0.16 [0.07–0.24] for PH, and 0.22 [0.15–0.29] for AUC. Bland-Altman plots showing these agreements are presented in Figure S16. Overall, the PSTH curve showed stronger intra-subject than inter-subject correlations across visits. However, the derived PT, PH, and AUC varied markedly from night to night. The results suggested that PT may be more state-like than trait-like.

## Supplemental discussion: Possible Mechanistic Speculations

Respiratory arousals are initiated when escalating respiratory drive crosses a threshold, thereby activating either cortical or subcortical pathways. These escalating stimuli typically arise from two sources: 1) chemical signals, such as increased arterial partial pressure of CO_2_ (hypercapnia) or lowered arterial partial pressure of O_2_ (hypoxia), and 2) mechanical signals reflecting growing inspiratory effort. Several factors modulate this arousal threshold (2), including the state of sleep itself. Slow wave sleep, including conventional N3 sleep, has a higher arousal threshold. Continuous sleep depth, as measured by the Odds Ratio Product, is a measure of arousability during sleep. The arousal threshold is low in the EEG state of cyclic alternating pattern (CAP) and high in non-CAP sleep. Medications such as benzodiazepines can increase the threshold by blunting the neural activities in the central nervous system. However, multivariable regression analysis revealed no significant correlations between PT and benzodiazepines (3), nor was there a direct correlation between PT and daytime sleepiness. Notably, obesity was strongly correlated with PT, potentially through an impact on upper airway collapsibility, which may delay mechanical airway reopening, a phenomenon commonly observed in OSA patients, as described below.

Arousal from sleep is a complex neurobiological phenomenon that occurs normally but may be amplified in frequency and intensity in disease. Components include the autonomic, respiratory, electrocortical, and, sometimes, motor systems. Awakening is controlled by a widely distributed network spanning multiple brain regions, including monoaminergic, cholinergic, glutamatergic, and peptidergic systems (e.g., cholinergic neurons in the pedunculopontine and laterodorsal tegmental nuclei and the basal forebrain, glutamatergic neurons in the central thalamus, orexin from the lateral hypothalamus, norepinephrine from the locus coeruleus). There is convergence of arousing stimuli onto brainstem neuronal groups, such as the lateral parabrachial complex (4). Efferent information moves through the basal forebrain and thalamus to cortical projections. The cumulative effect of arousals over time could damage this network, which also mediates numerous critical autonomic functions. In this context, a longer PT may indicate a slower rate of propagation of the brainstem-cortical network. However, the prolongation of PT is likely due not solely to structural slowing but also to functional adaptation. There is emerging evidence that arousals decrease with untreated OSA. This “blunting” of arousals may represent an adaptive phenomenon or be secondary to the adverse effects of chronic hypoxemia on the aforementioned arousal centers (5–7).

---

**Figure S1.**
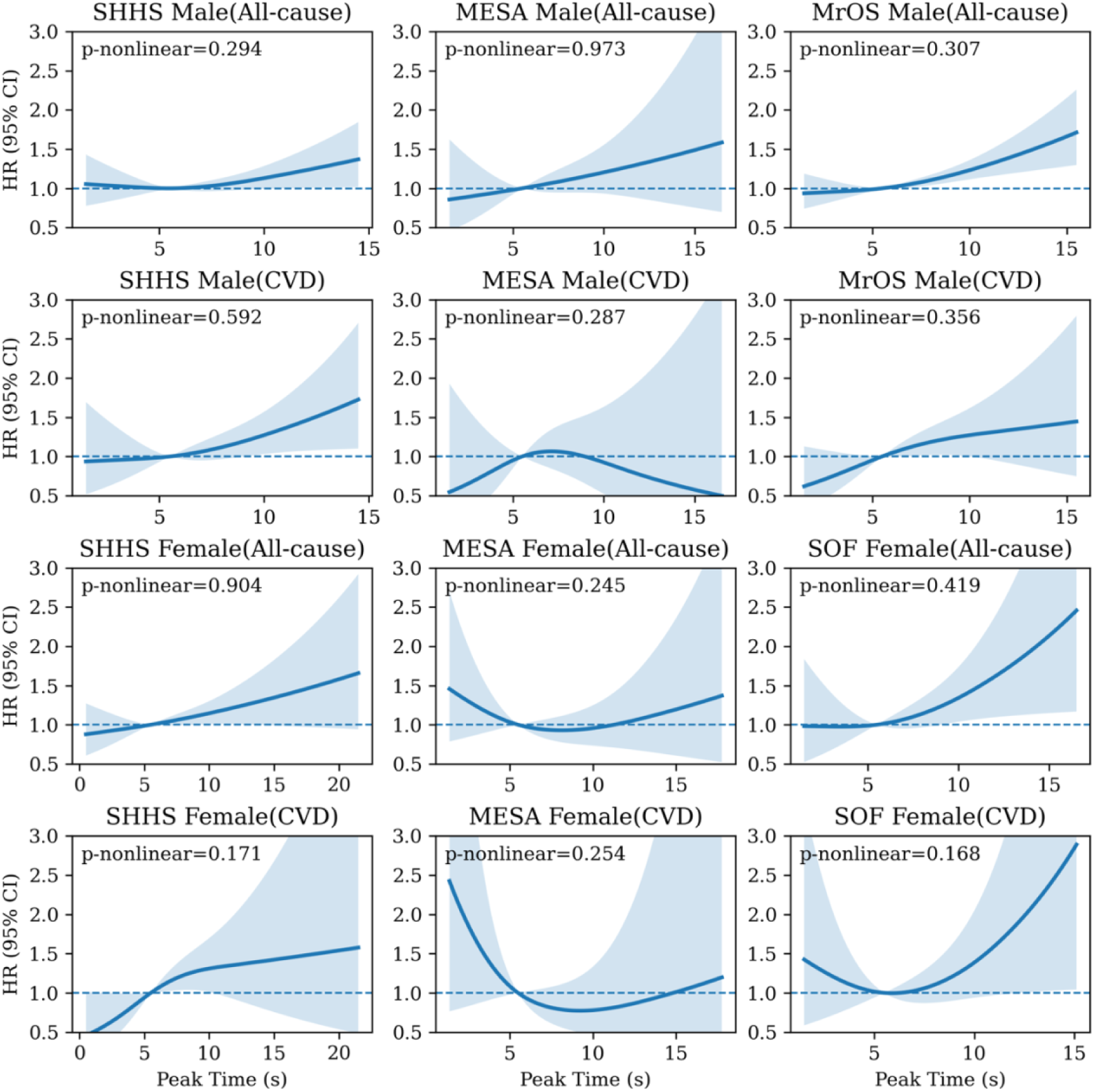
Restricted cubic spline assessment of linearity for Peak Time (PT). PT was modeled using a 3-knot restricted cubic spline with knots at the 5th, 50th, and 95th percentiles, with hazard ratios referenced to the median PT. Evidence for nonlinearity was evaluated using a likelihood ratio test comparing the spline model to the linear PT model p-nonlinear). Across cohorts and outcomes, p-nonlinear values provided little evidence of departure from linearity, supporting the linear PT specification used in the primary analyses.

**Figure S2.**
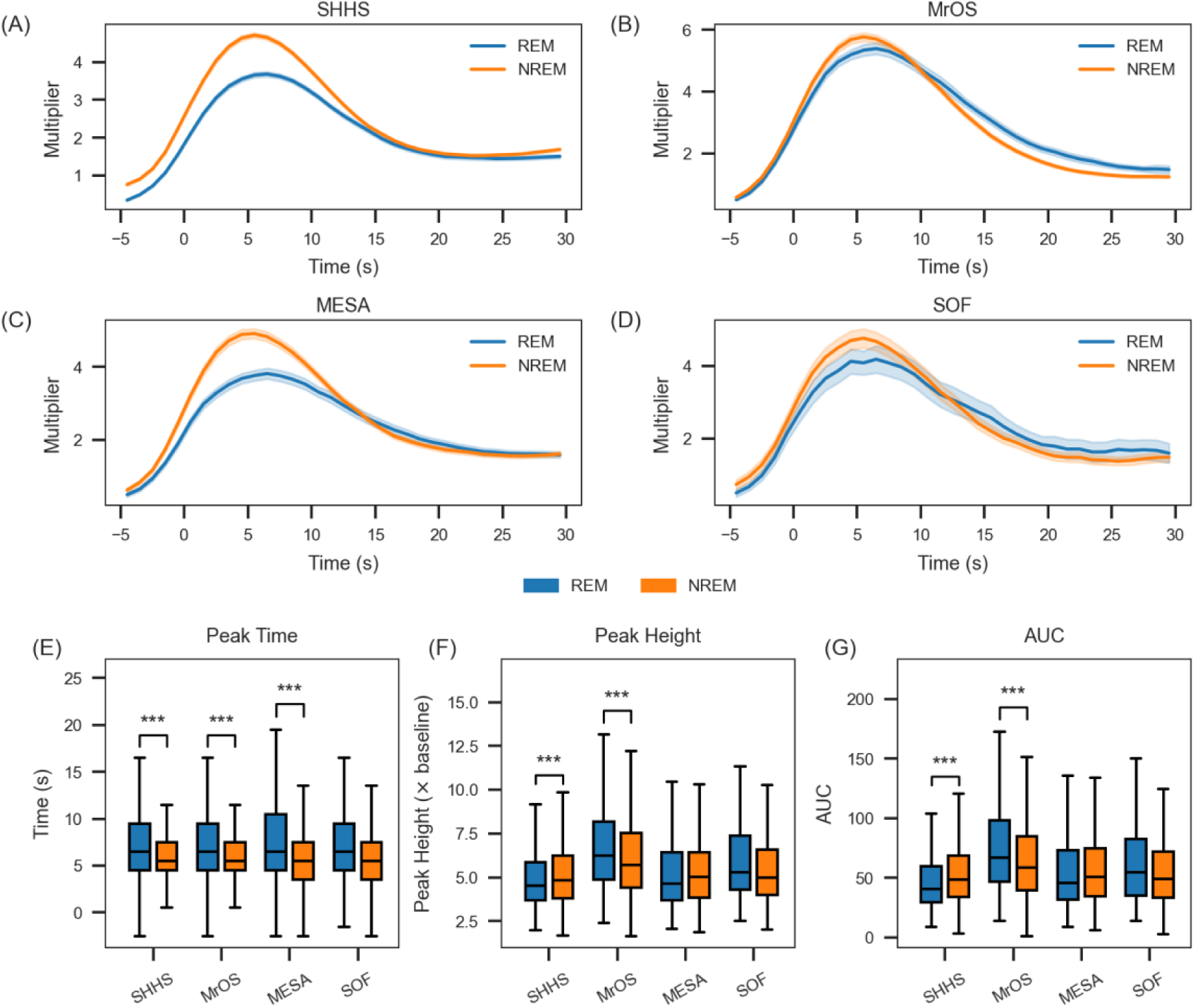
Peri-Stimulus Time Histograms (PSTHs) and the derived features stratified by REM vs. NREM sleep (A) SHHS, (B) MrOS, (C) MESA, and (D) SOF. In each panel, the solid lines show the average arousal rate for REM (blue) and NREM (orange), with shaded regions representing 95% confidence intervals. (E)-(G): Box plots compare peak time (PT), peak height (PH) and AUC between REM (blue) and NREM (orange) in each cohort. Across all cohorts, PSTH curves and PT values differ by sleep stage, with NREM generally exhibiting a higher PH and an earlier PT than REM. **\***: p < 0.05; **: p<0.01; ***: p<0.001.

**Figure S3.**
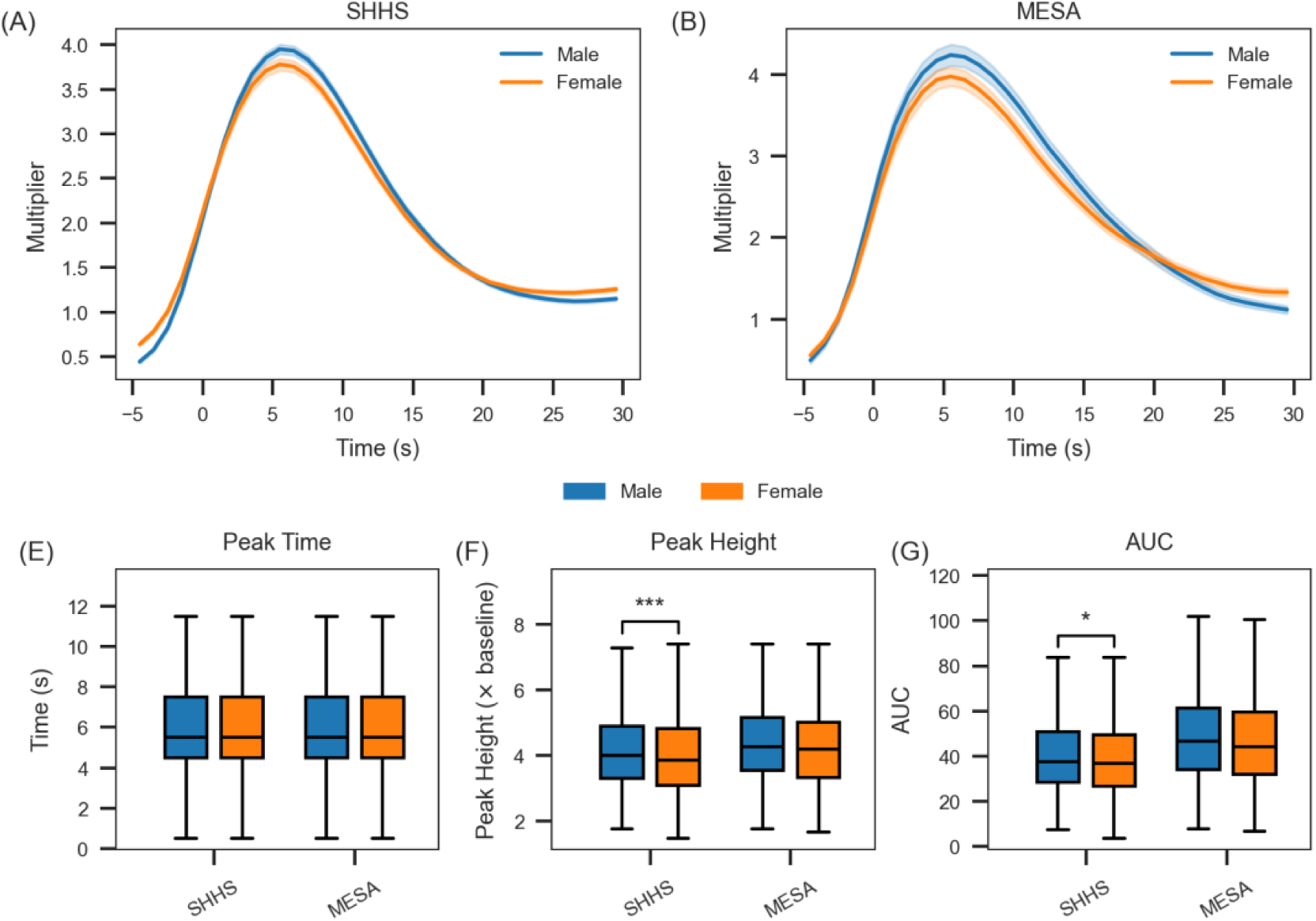
Sex-Specific PSTHs and Feature Comparisons (A) and (B): Population-averaged PSTH curves, with males (blue) and females (orange) plotted separately; shaded areas represent approximate confidence intervals. In both cohorts, males exhibit a higher arousal rate at the peak than females, though the shape and timing of the peak differ slightly across cohorts. (C)-(G): Box plots of PT, PH, and AUC stratified by sex. In SHHS, males exhibited slightly higher PH (p <0.001, Cohen’s d=0.13) and AUC (p = 0.04, Cohen’s d = 0.04) than females. However, the effect size was negligible. *: p < 0.05; **: p<0.01; ***: p<0.001.

**Figure S4.**
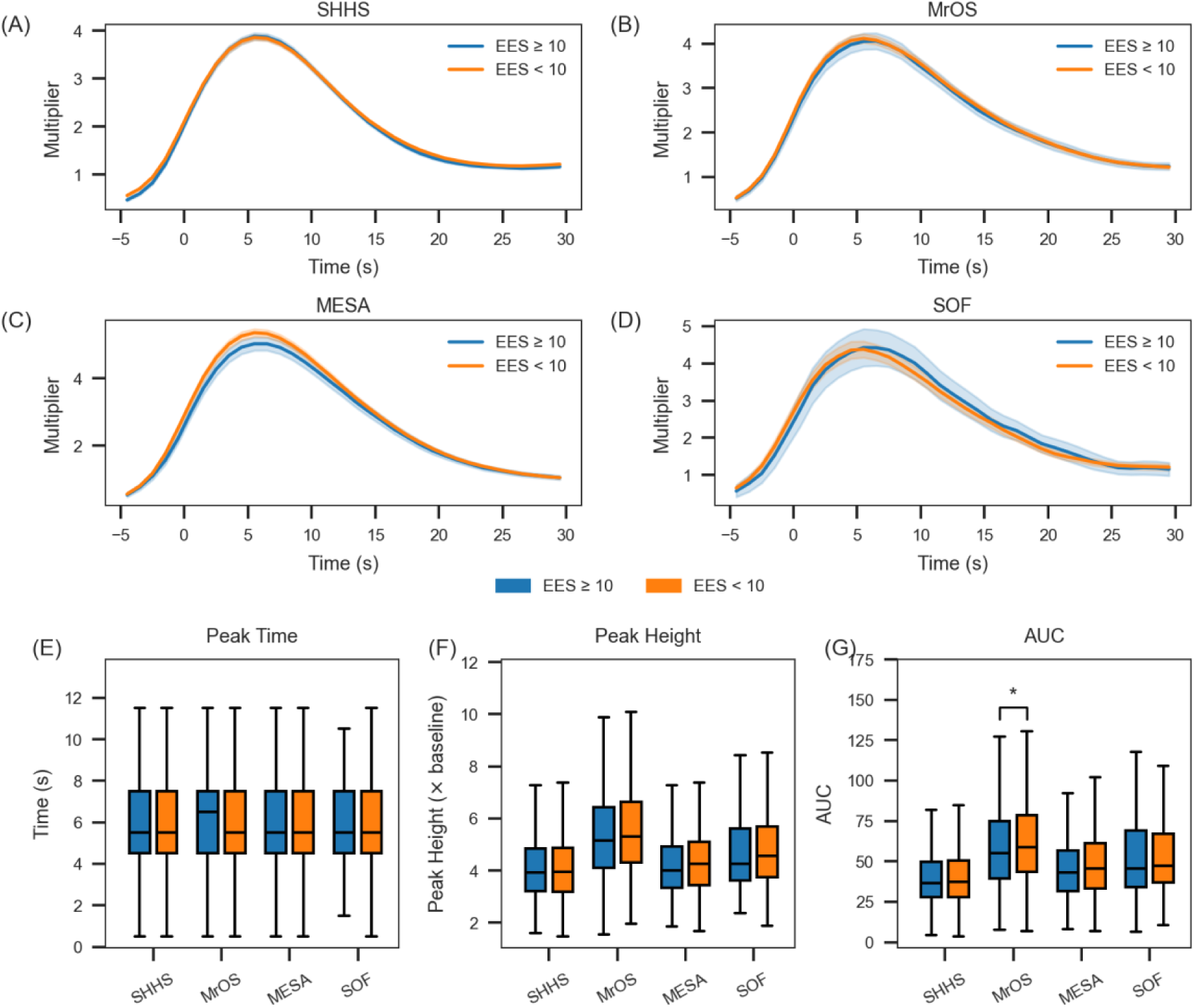
PSTHs and Features by Daytime Sleepiness (ESS ≥10 vs <10) (A) – (D): Mean PSTH curves stratified by participants with ESS ≥ 10 (blue) vs. ESS < 10 (orange). Shaded regions represent approximate 95% confidence intervals. (E)-(G): Box plots of peak time, peak height and AUC across cohort by ESS group. PSTH of different ESS groups largely overlapped. In MrOS, the AUC feature showed a significant difference (p=0.033, Cohen’s d =-0.15) between ESS groups.

**Figure S5.**
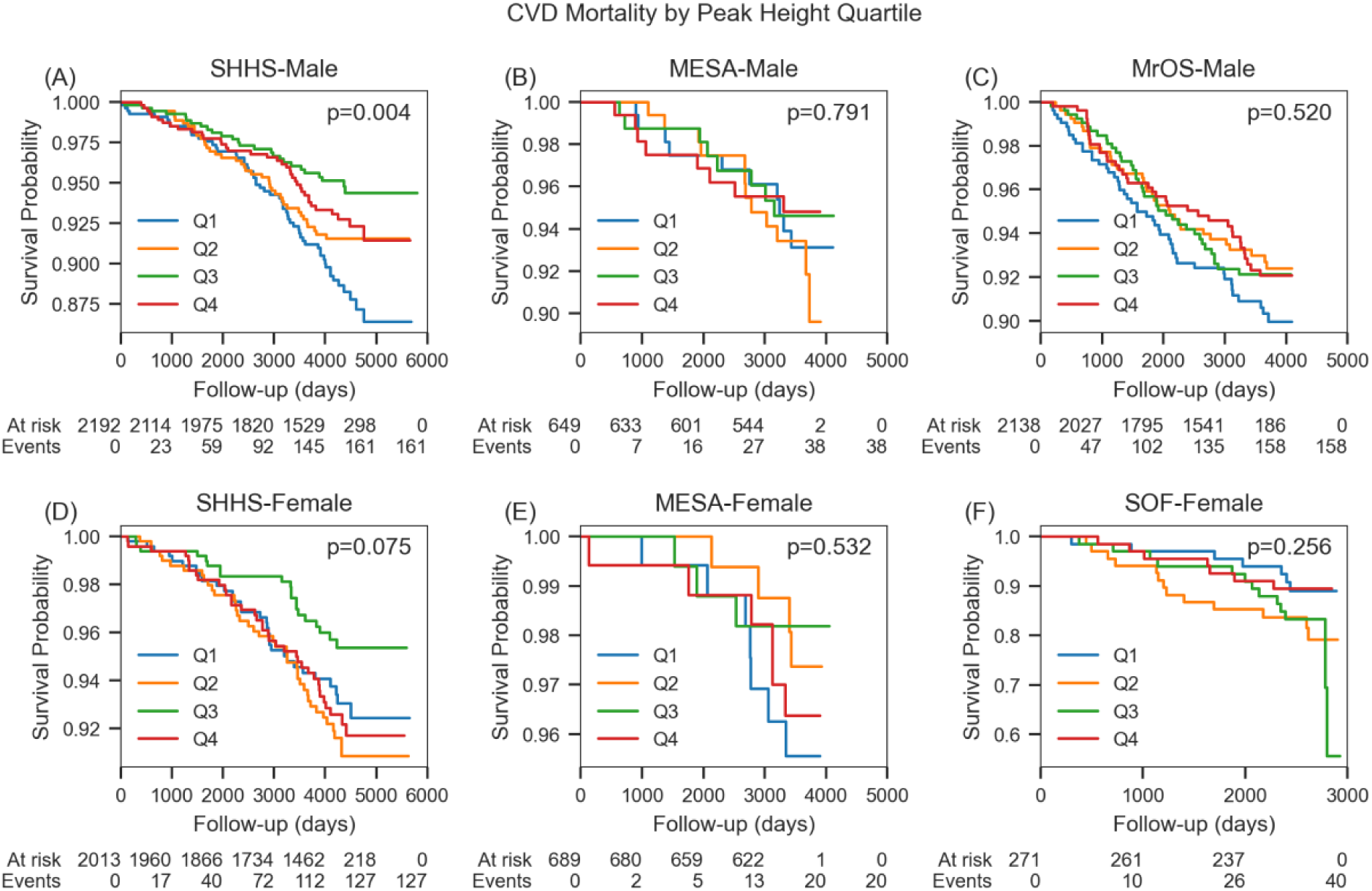
CVD Mortality Kaplan-Meier Curves by PH Quartiles. Panels (A)–(C) correspond to males (SHHS, MESA, MrOS), and panels (D)–(F) to females (SHHS, MESA, SOF). The x-axis indicates follow-up time (days), and the y-axis depicts cardiovascular event-free survival. Numbers below each panel show participants at risk (“At risk”) and the cumulative number of events (“Events”) at designated intervals. Log-rank tests were performed to compare survival differences among the four PH quartiles, with p-values displayed in the top-right corners of each plot.

**Figure S6.**
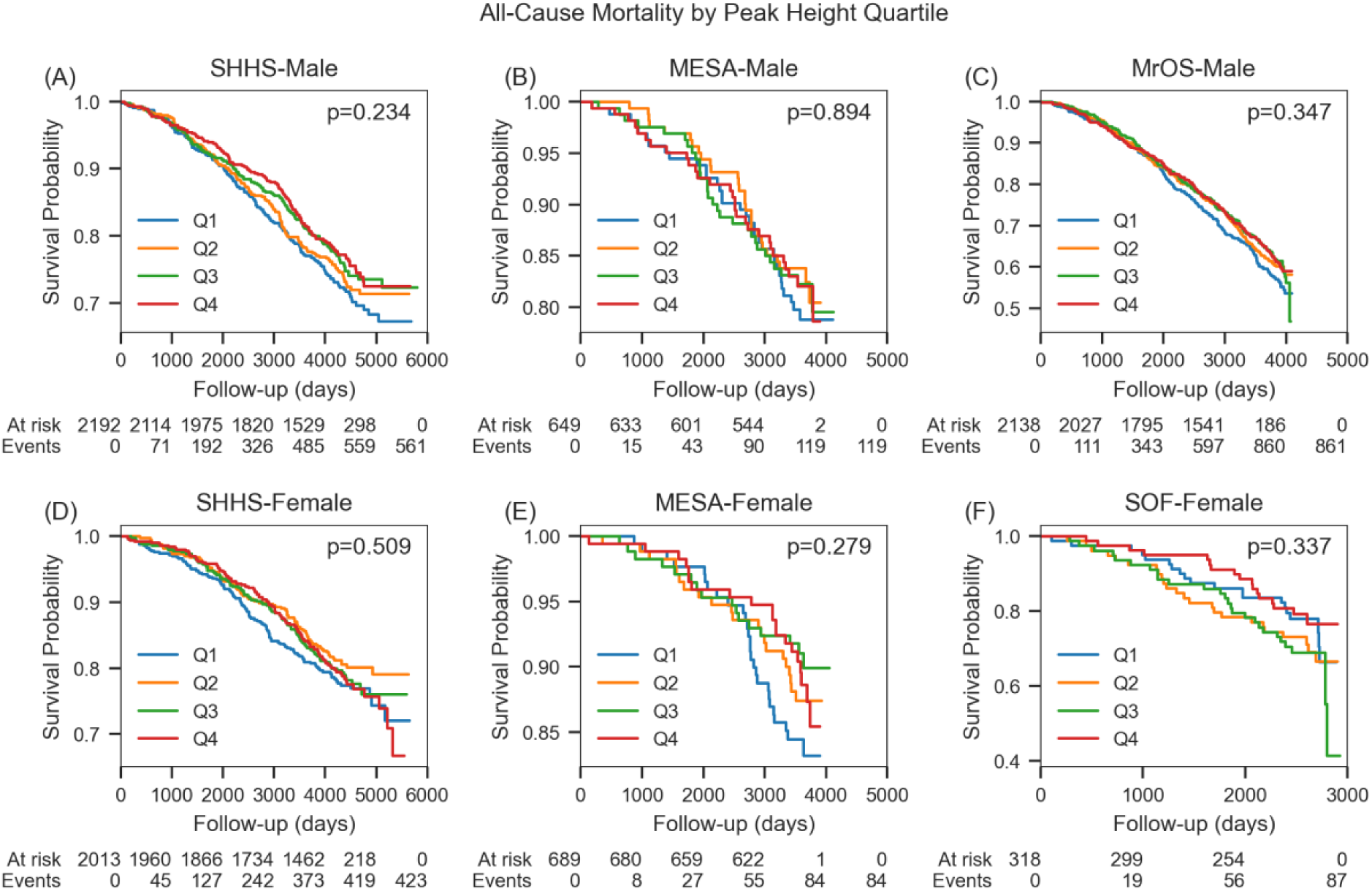
All-Cause Mortality Kaplan-Meier Curves by PH Quartiles Panels (A)–(C) correspond to males (SHHS, MESA, MrOS), and panels (D)–(F) to females (SHHS, MESA, SOF). The x-axis indicates follow-up time (days), and the y-axis depicts the probability of survival from any cause. Numbers below each panel show participants at risk (“At risk”) and the cumulative number of events (“Events”) at designated intervals. Log-rank tests were performed to compare survival differences among the four PH quartiles, with p-values displayed in the top-right corners of each plot.

**Figure S7.**
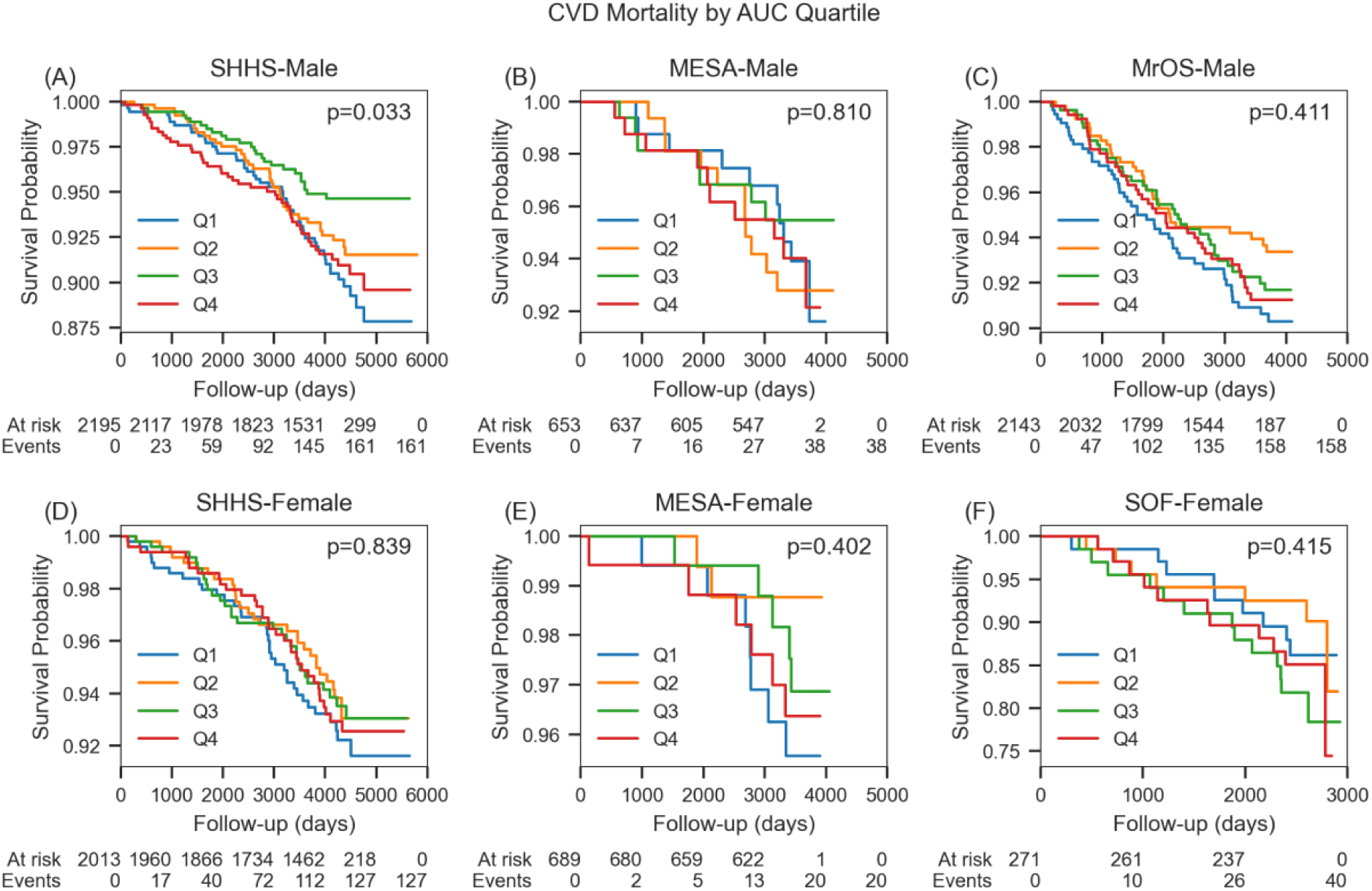
CVD Mortality Kaplan-Meier Curves by AUC Quartiles Panels (A)–(C) correspond to males (SHHS, MESA, MrOS), and panels (D)–(F) to females (SHHS, MESA, SOF). The x-axis indicates follow-up time (days), and the y-axis depicts cardiovascular event-free survival. Numbers below each panel show participants at risk (“At risk”) and the cumulative number of events (“Events”) at designated intervals. Log-rank tests were performed to compare survival differences among the four AUC quartiles, with p-values displayed in the top-right corners of each plot.

**Figure S8.**
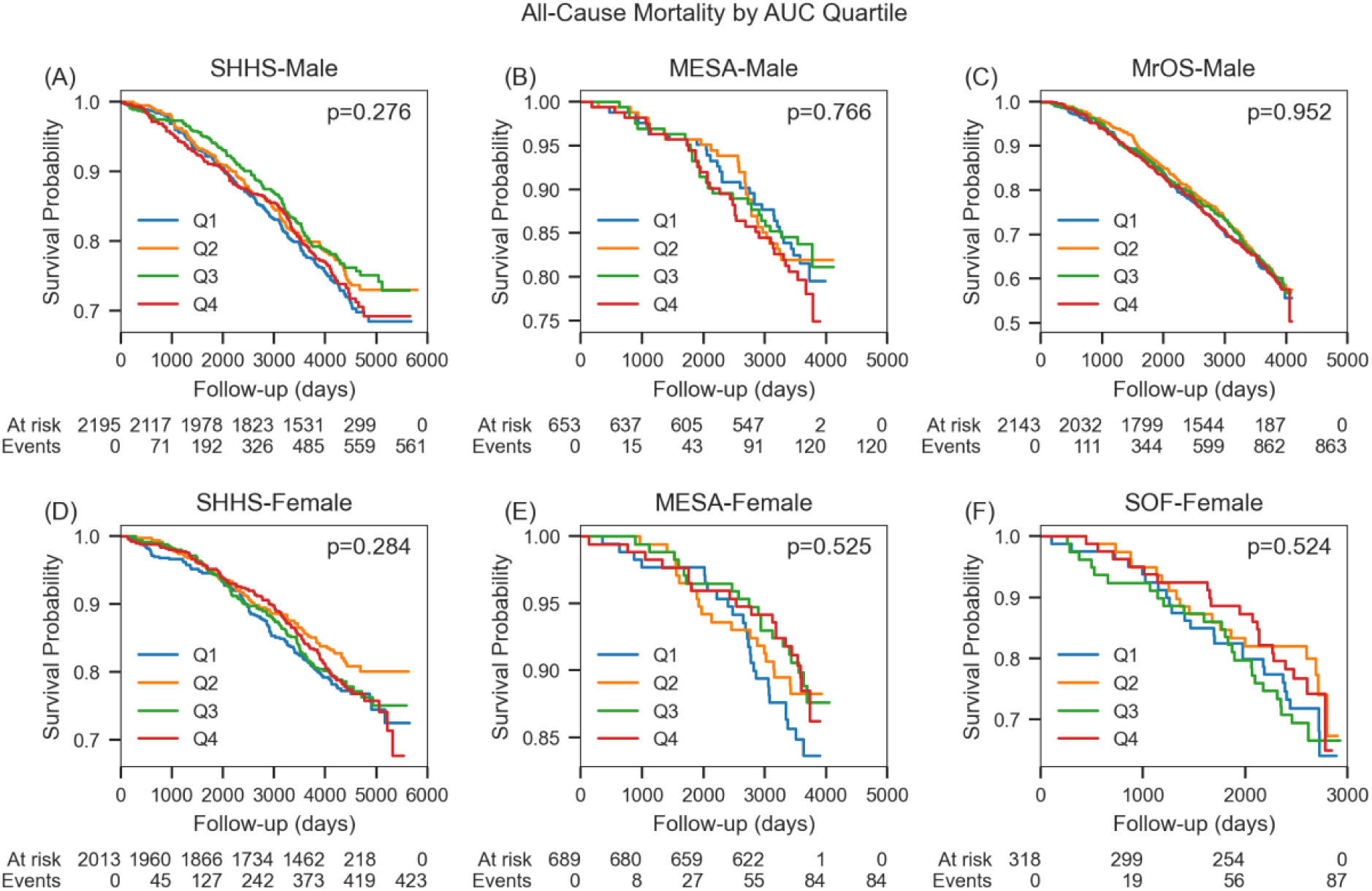
All-Cause Mortality Kaplan-Meier Curves by AUC Quartiles. Panels (A)–(C) correspond to males (SHHS, MESA, MrOS), and panels (D)–(F) to females (SHHS, MESA, SOF). The x-axis indicates follow-up time (days), and the y-axis depicts the probability of survival from any cause. Numbers below each panel show participants at risk (“At risk”) and the cumulative number of events (“Events”) at designated intervals. Log-rank tests were performed to compare survival differences among the four AUC quartiles, with p-values displayed in the top-right corners of each plot.

**Figure S9.**
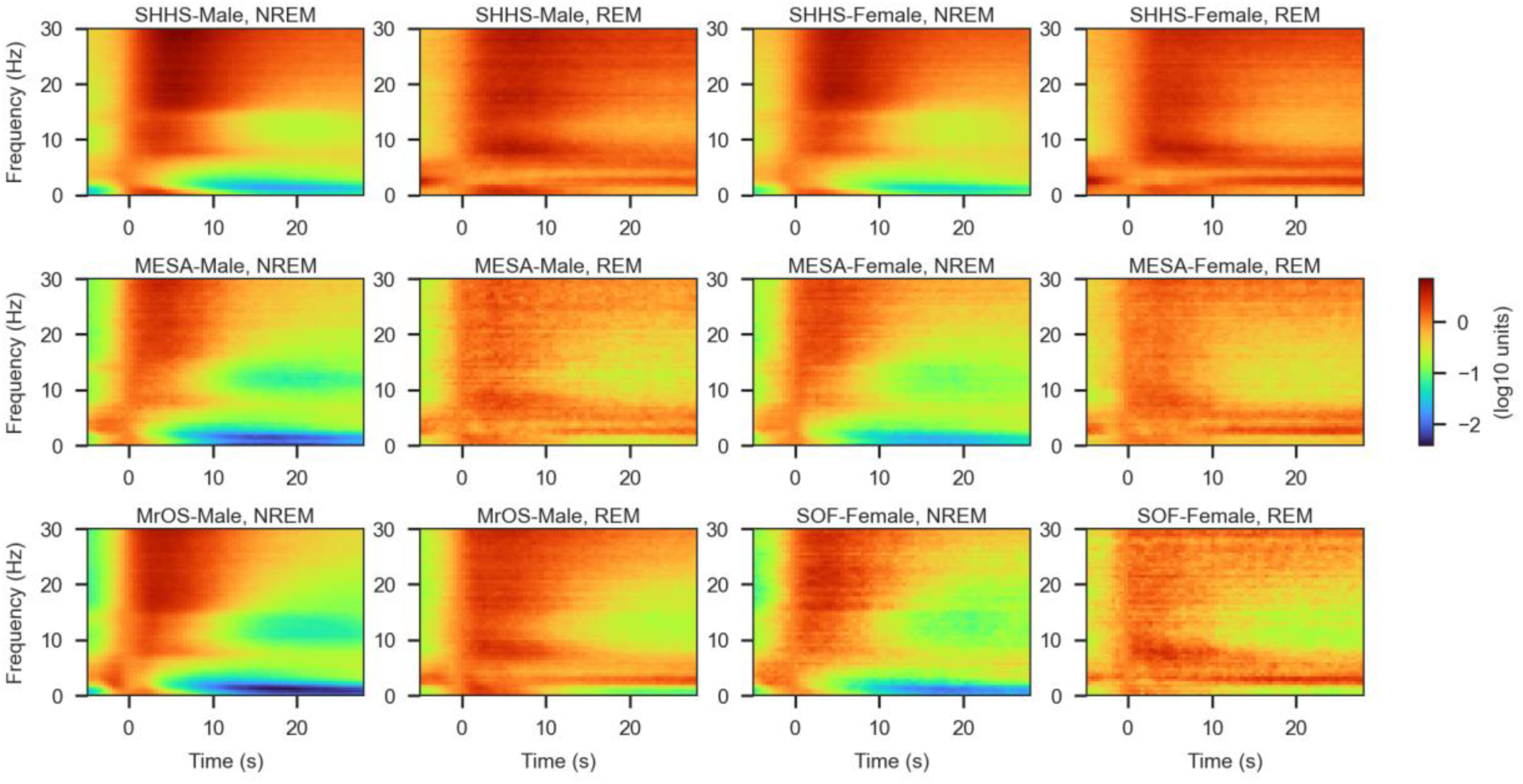
EEG Time–Frequency Spectrum Aligned to Apnea Termination. EEG spectrums were aligned to the end of each apnea event (t=0 s). Spectra were estimated with Welch’s method (2 s Hanning window, 75 % overlap) from the C3–A2 derivation, except for the MESA cohort, which used C4–A1. Power at every time–frequency bin was log-transformed, normalized to the power at t = 0 s, and then averaged within sex and sleep stage (REM vs. NREM). In all panels, a surge of high-frequency power is evident immediately after apnea termination.

**Figure S10.**
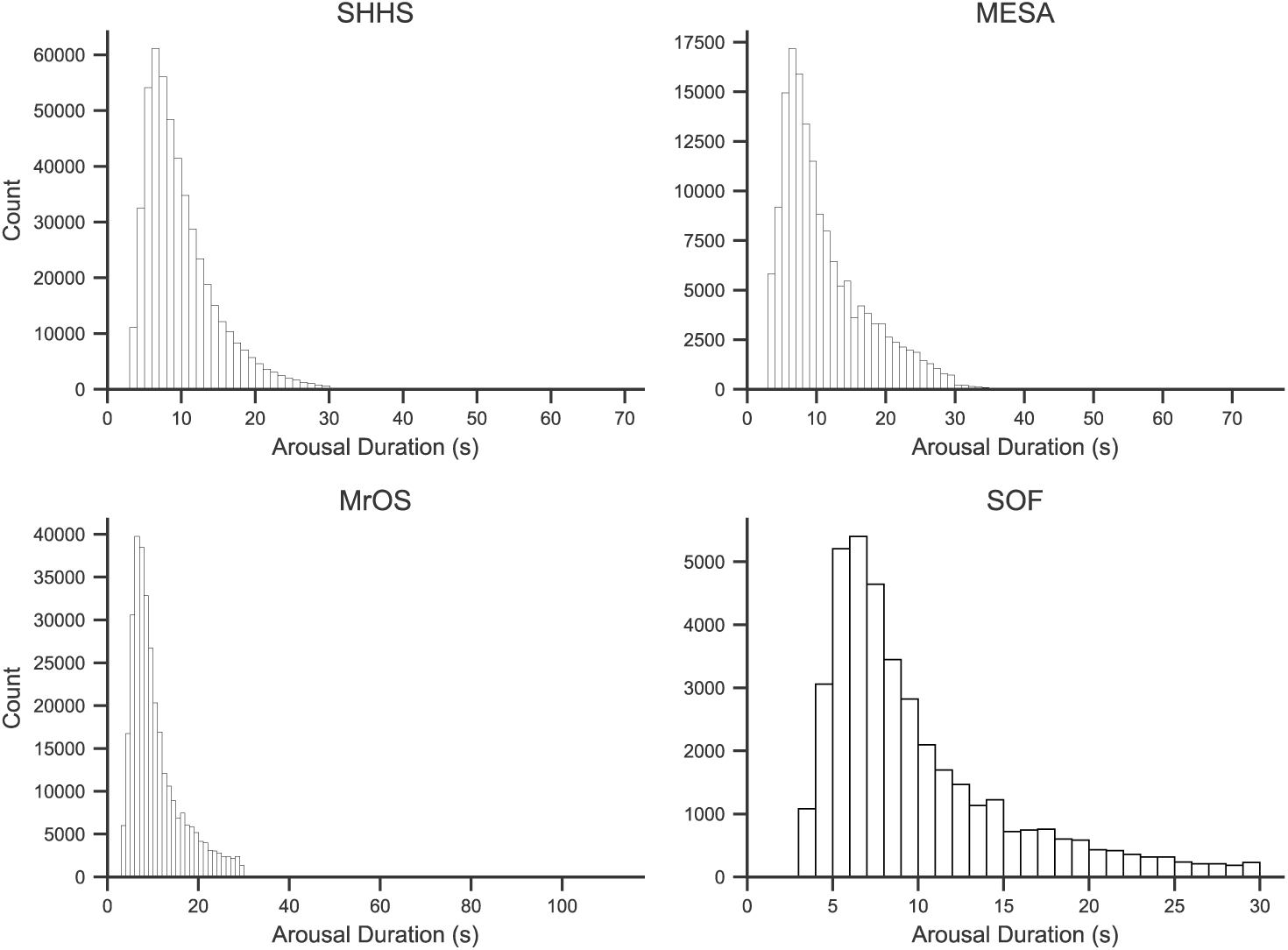
Distribution of Arousal Event Duration Across Cohorts

**Figure S11.**
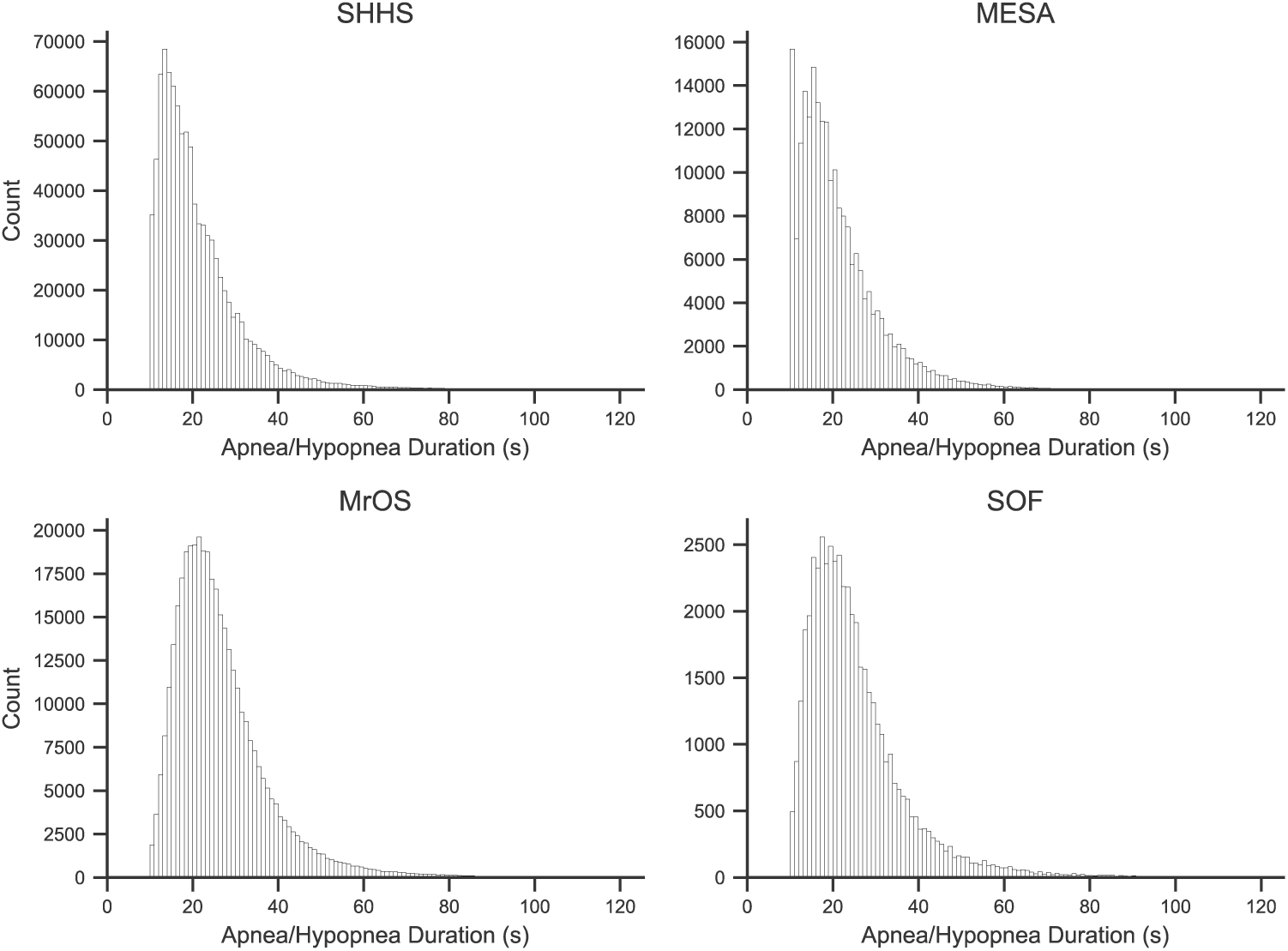
Distribution of Apnea/Hypopnea Event Duration Across Cohorts

**Figure S12.**
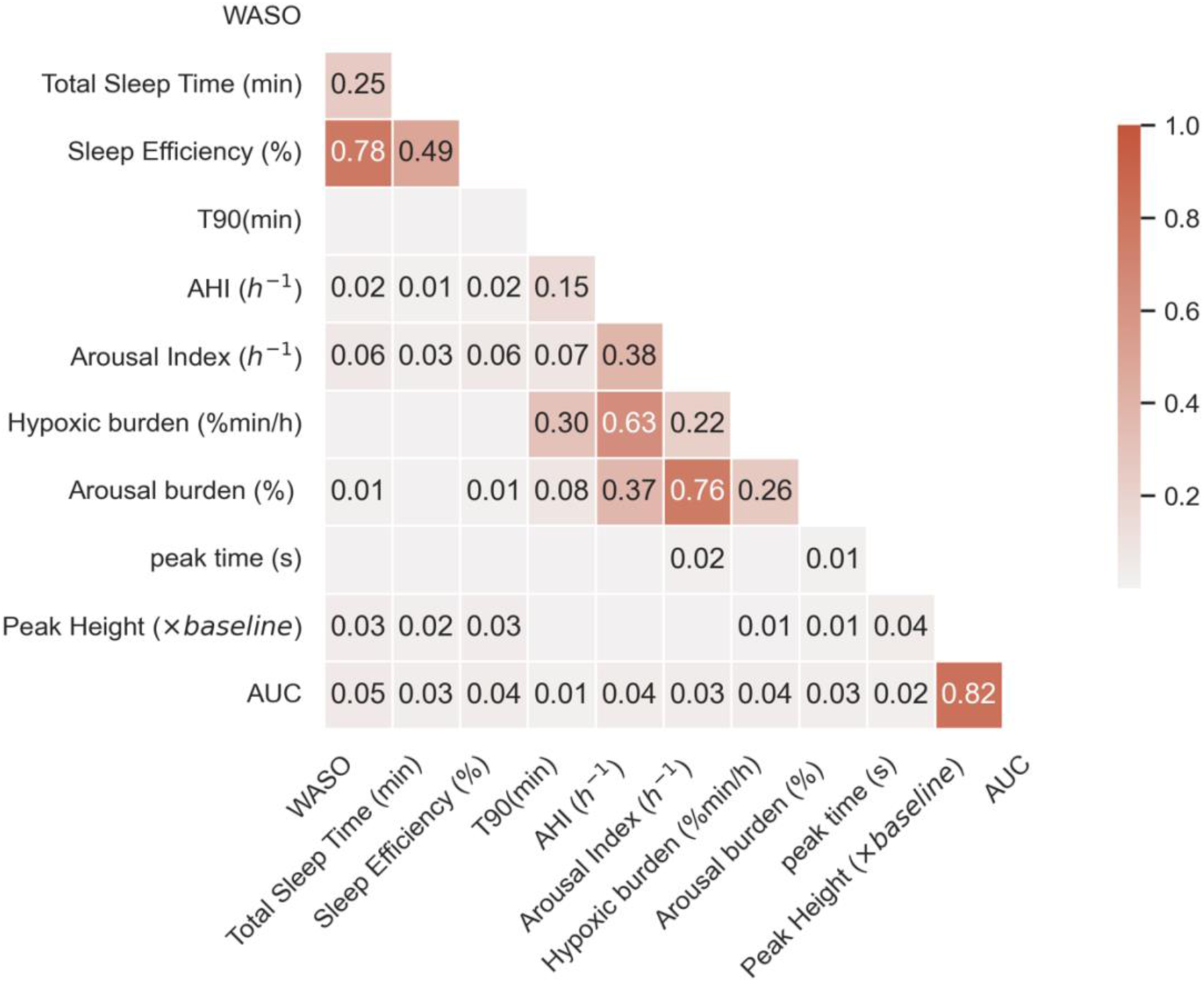
Correlation (R²) Between PSTH Features and Conventional Sleep Metrics Relationship (R²) between PSTH features (peak time, peak heigh and the AUC above baseline), and conventional sleep metrics (AHI, arousal index, hypoxic burden, etc.). The low R² values indicate that PT accounts for only a small portion of the variance in these measures, suggesting it captures a distinct physiologic dimension of sleep-disordered breathing.

**Figure S13.**
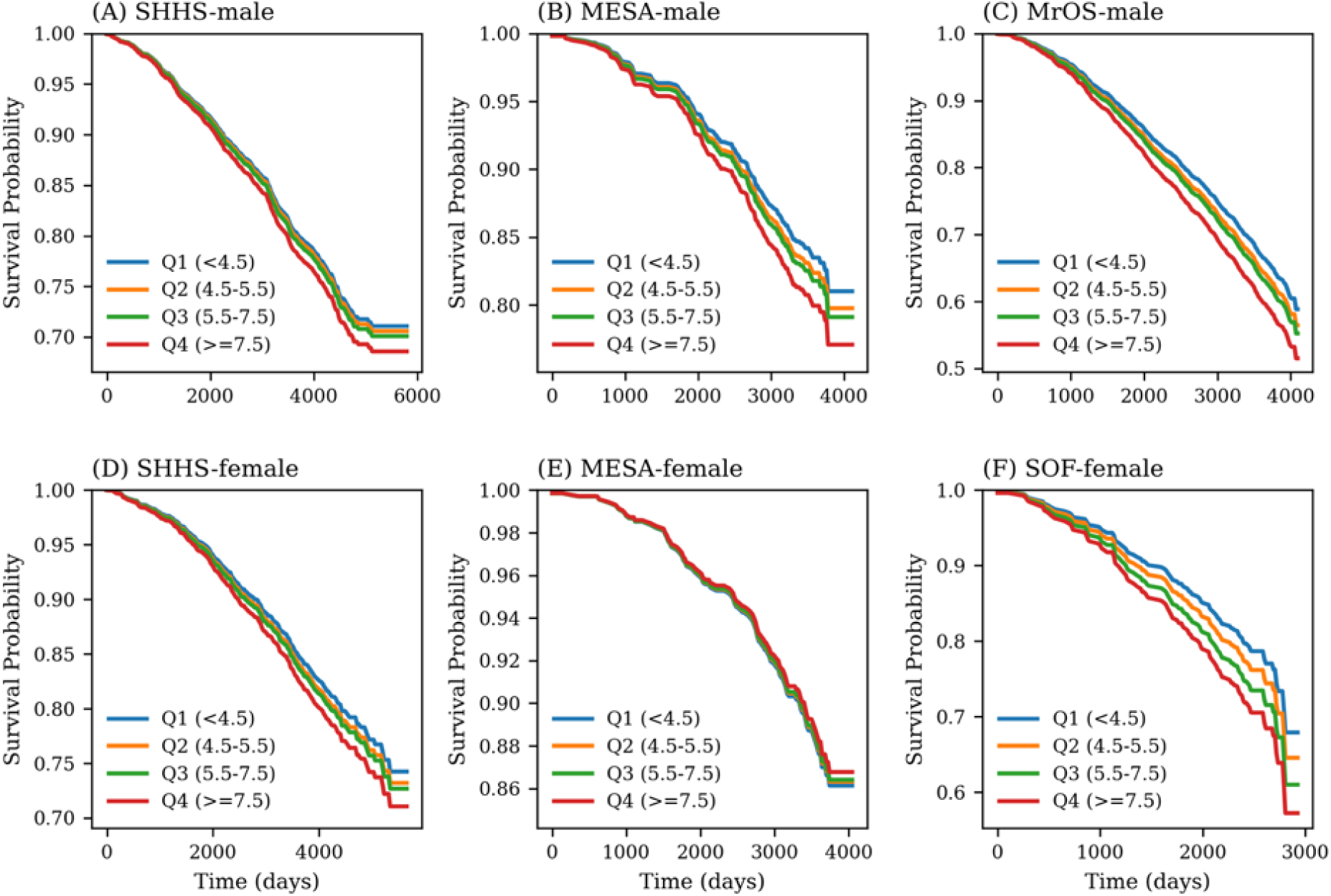
Adjusted Survival Curves from Cox Model by PT The models incorporating peak time (PT) as a continuous predictor. For illustrative purposes, separate curves are shown for several PT cutoffs (e.g., < 4.5 s, 4.5–5.5 s, 5.5–7.5 s, and ≥ 7.5 s). These curves reflect estimated survival probabilities from the fitted model rather than direct Kaplan-Meier curves, illustrating how survival differs across PT categories. The x-axis represents follow-up time (days), and the y-axis shows the estimated survival probability.

**Figure S14.**
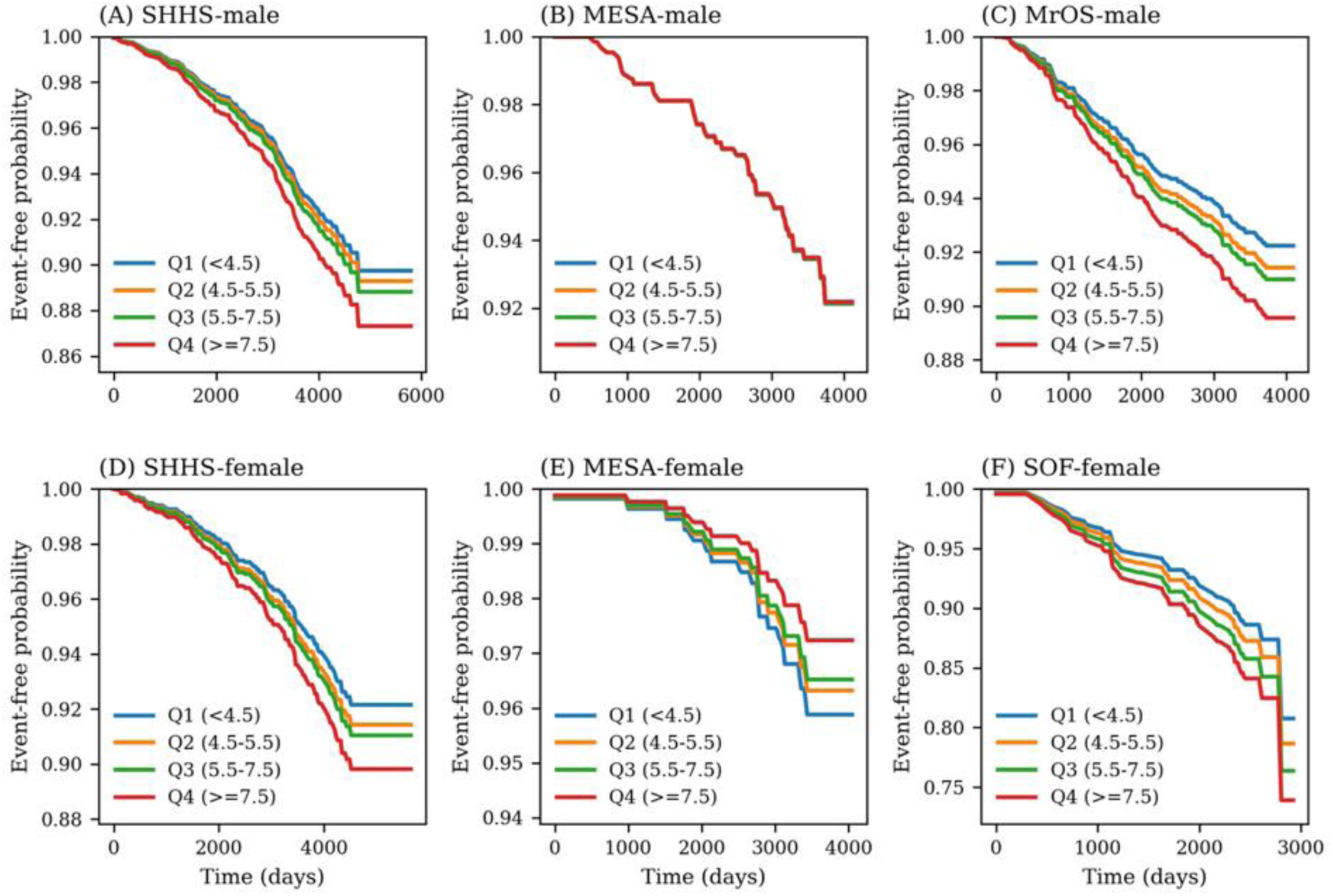
Adjusted CVD Event-Free Curves from Cox Model by PT The models incorporating peak time (PT) as a continuous predictor. For illustrative purposes, separate curves are shown for several PT cutoffs (e.g., < 4.5 s, 4.5–5.5 s, 5.5–7.5 s, and ≥ 7.5 s). These curves reflect estimated survival probabilities from the fitted model rather than direct Kaplan-Meier curves, illustrating how survival differs across PT categories. The x-axis represents follow-up time (days), and the y-axis shows the estimated survival probability.

**Figure S15.**
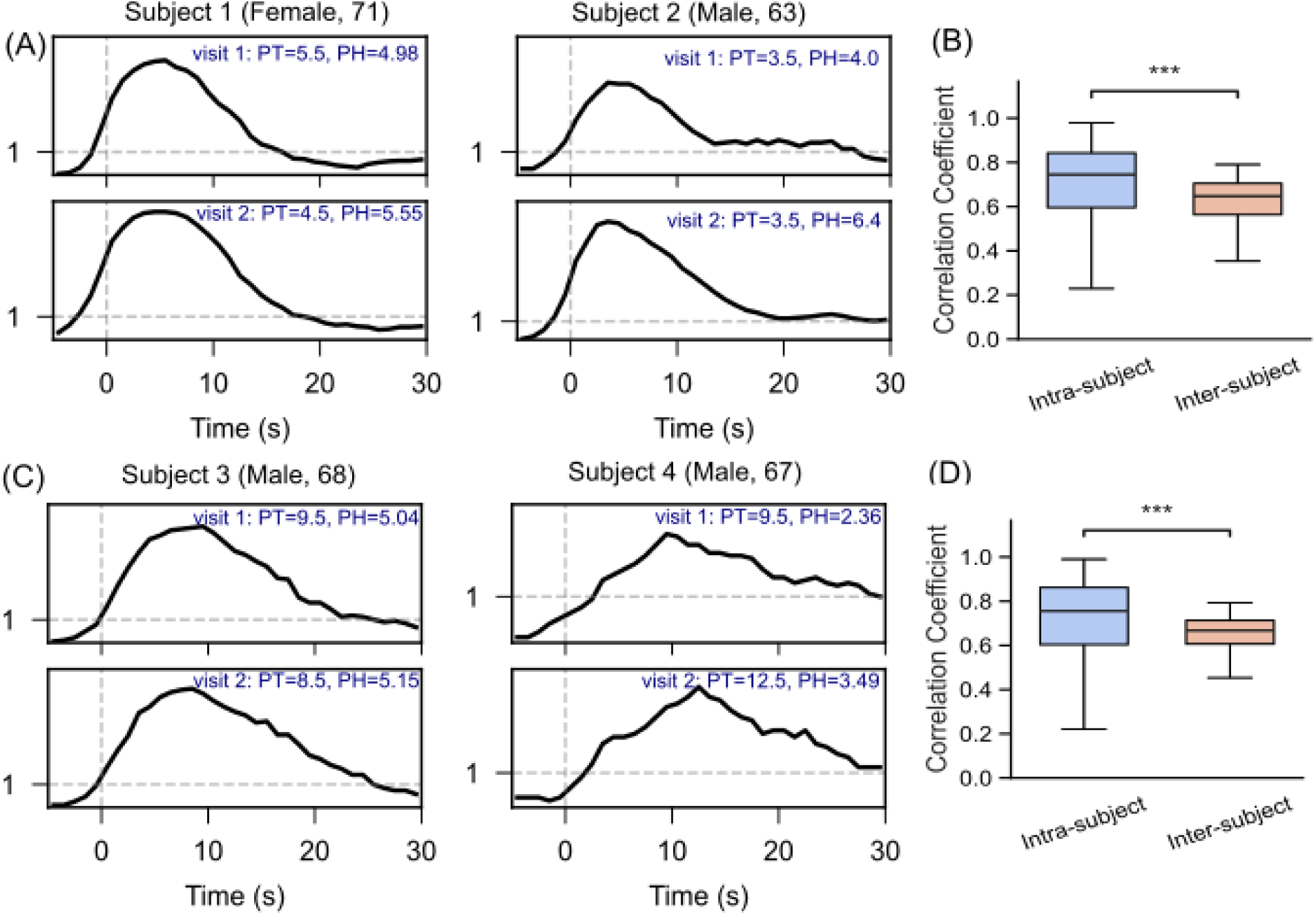
Within-Subject PSTH Consistency Across Visits (SHHS and MrOS). (A) Representative PSTHs from four participants (labeled by sex and age) who underwent repeated polysomnography recordings. Each subplot displays PSTHs for Visit 1 and Visit 2, along with the corresponding PT and PH. Although the PSTH shape tends to remain similar, PT, PH, and AUC shift between visits. (B) Box plots of Pearson correlation coefficients for PSTH curves show higher intra-subject than inter-subject correlation (p < 0.001). Top: SHHS; Bottom: MrOS.

**Figure S16.**
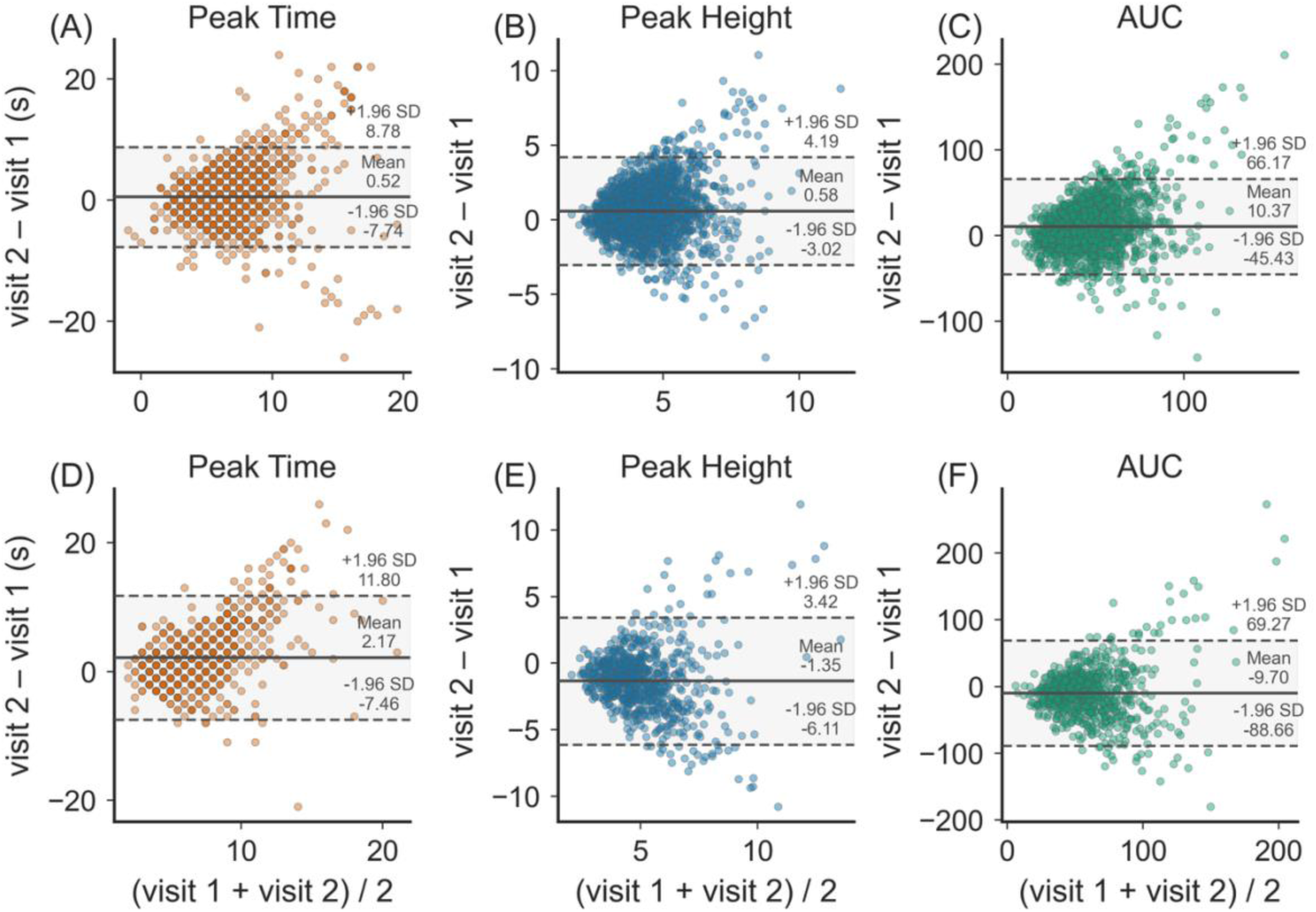
Bland-Altman Plots of Test-Retest Reproducibility for PSTH Features Bland-Altman plots assessing the test-retest reproducibility of PSTH features in SHHS (top) and MrOS (bottom). In each plot, the y-axis represents the difference between the features derived from two visits (Visit 2 – Visit 1), and the x-axis represents the average of the features from two visits. The solid central line indicated the mean differences, while the dashed outer lines represent the 95% limits of agreement (mean±1.96 SD).

**Table S1.** Stage-Specific PT (REM/NREM) and All-Cause/CVD Mortality.

| Group | PT-REM |  |  |  | PT-NREM |  |  |  |
| --- | --- | --- | --- | --- | --- | --- | --- | --- |
|  | All-cause mortality |  | CVD mortality |  | All-cause mortality |  | CVD mortality |  |
|  | HR<br>(95%CI) | P-value | HR<br>(95%CI) | P-value | HR<br>(95%CI) | P-value | HR<br>(95%CI) | P-value |
| SHHS-Male | 1.00<br>(0.97-1.02) | 0.85 | 1.01<br>(0.97-1.05) | 0.68 | 1.01<br>(0.99-1.04) | 0.31 | 1.01<br>(0.97-1.07) | 0.56 |
| SHHS-Female | 1.00<br>(0.97-1.02) | 0.83 | 0.98<br>(0.94-1.03) | 0.45 | **1.03**<br>**(1.01-1.06)** | **0.006** | **1.06**<br>**(1.02-1.11)** | **0.006** |
| SOF-Female | 1.00<br>(0.92-1.07) | 0.90 | 1.04<br>(0.90-1.21) | 0.60 | 1.02<br>(0.97-1.07) | 0.44 | 1.07<br>(0.99-1.15) | 0.074 |
| MrOS-Male | 0.99<br>(0.97-1.02) | 0.60 | 1.02<br>(0.97-1.07) | 0.41 | **1.04**<br>**(1.02-1.06)** | **<0.001** | 1.02<br>(0.97-1.07) | 0.41 |

Hazard ratios (HRs) and 95% confidence intervals are shown for a 1-second increase in PT; p-values reflect Wald tests within Cox proportional hazards models. Models were adjusted for demographic factors (age, race), body mass index, and comorbidities (hypertension, diabetes, smoking, depression, stroke). PT-REM and PT-NREM were analyzed in distinct models to assess whether delayed arousal peak time in each sleep stage independently predicts mortality risk.

**Table S2.** Covariates Associated with Peak Time (PT) in SHHS.

| Variables | All |  | Male |  | Female |  |
| --- | --- | --- | --- | --- | --- | --- |
| | $\beta$ (95% CI) | p | $\beta$ (95% CI) | p | $\beta$ (95% CI) | p |
| Age | **0.02 (0.02, 0.03)** | **<0.001** | **0.03 (0.02, 0.04)** | **<0.001** | **0.02 (0.00, 0.03)** | **0.028** |
| Female Sex | -0.10 (-0.28, 0.09) | 0.320 | - | - | - | - |
| BMI | **0.04 (0.02, 0.06)** | **<0.001** | **0.04 (0.02, 0.07)** | **0.001** | **0.04 (0.01, 0.07)** | **0.005** |
| Benzodiazepine | -0.06 (-0.48, 0.37) | 0.796 | 0.21 (-0.36, 0.79) | 0.467 | -0.21 (-0.82, 0.41) | 0.511 |
| CVD History | **0.42 (0.18, 0.67)** | **<0.001** | **0.41 (0.14, 0.68)** | **0.003** | 0.39 (-0.05, 0.84) | 0.084 |
| COPD | -0.51 (-1.45, 0.43) | 0.290 | -0.87 (-1.99, 0.25) | 0.126 | -0.12 (-1.68, 1.44) | 0.878 |
| MAP | 0.004 (-0.00, 0.01) | 0.352 | 0.003 (-0.01, 0.01) | 0.450 | 0.004 (-0.01, 0.02) | 0.481 |
| AHI | **-0.02 (-0.02, -0.01)** | **<0.001** | **-0.01 (-0.02, -0.01)** | **<0.001** | **-0.03 (-0.04, -0.01)** | **<0.001** |

Coefficients ( $\beta$ ) and 95% confidence intervals (CI) are derived from linear regression models where PT is the dependent variable, statistically significant results ( $p < 0.05$ ) indicate covariates that drive higher or lower PT, adjusting for the other listed covariates. BMI, body mass index; AHI, apnea-hypopnea index; MAP, mean arterial pressure; CVD, cardiovascular disease; COPD, chronic obstructive pulmonary disease. "Gender (Female)" denotes a binary variable comparing females to the reference (male) group. $\beta$ represents the average change in PT per 1-unit increase in the continuous covariate or the mean difference relative to the reference group for categorical variables. Values in bold indicate a significant association, $p < 0.05$ .

**Table S3.** Covariates Associated with Peak Height (PH) in SHHS.

| Variables | All |  | Male |  | Female |  |
| --- | --- | --- | --- | --- | --- | --- |
| | $\beta$ (95% CI) | p | $\beta$ (95% CI) | p | $\beta$ (95% CI) | p |
| Age | 0.004 (-0.00, 0.01) | 0.086 | **-0.004 (-0.01, 0.00)** | **0.208** | 0.01 (0.00, 0.02) | **0.001** |
| Female Sex | **-0.12 (-0.20, -0.03)** | **0.010** | - | - | - | - |
| BMI | **-0.02 (-0.03, -0.01)** | **<0.001** | **-0.02 (-0.03, -0.00)** | **0.022** | -0.02 (-0.03, -0.01) | **<0.001** |
| Benzodiazepine | -0.002 (-0.20, 0.20) | 0.987 | -0.15 (-0.44, 0.14) | 0.321 | 0.08 (-0.20, 0.35) | 0.582 |
| CVD History | -0.01 (-0.12, 0.11) | 0.885 | 0.01 (-0.13, 0.14) | 0.936 | 0.002 (-0.20, 0.20) | 0.982 |
| COPD | -0.15 (-0.59, 0.29) | 0.508 | 0.13 (-0.43, 0.70) | 0.638 | -0.48 (-1.17, 0.22) | 0.180 |
| MAP | -0.002 (-0.00, 0.00) | 0.389 | -0.004 (-0.01, 0.00) | 0.066 | 0.001 (-0.00, 0.01) | 0.807 |
| AHI | **-0.004 (-0.01, -0.00)** | **0.006** | -0.005 (-0.01, -0.00) | **0.006** | -0.003 (-0.01, 0.00) | 0.217 |

Coefficients ( $\beta$ ) and 95% confidence intervals (CI) are derived from linear regression models where peak height (PH) is the dependent variable, statistically significant results ( $p < 0.05$ ) indicate covariates that drive higher or lower PH, adjusting for the other listed covariates. BMI, body mass index; AHI, apnea-hypopnea index; MAP, mean arterial pressure; CVD, cardiovascular disease; COPD, chronic obstructive pulmonary disease. "Gender (Female)" denotes a binary variable comparing females to the reference (male) group. $\beta$ represents the average change in PH per 1-unit increase in the continuous covariate or the mean difference relative to the reference group for categorical variables. Values in bold indicates a significant association, $p < 0.05$ .

**Table S4.** Covariates Associated with AUC Above Baseline in SHHS.

| Variables | All |  | Male |  | Female |  |
| --- | --- | --- | --- | --- | --- | --- |
| | $\beta$ (95% CI) | p | $\beta$ (95% CI) | p | $\beta$ (95% CI) | p |
| Age | **0.10 (0.04, 0.15)** | **0.001** | 0.03 (-0.04, 0.11) | 0.372 | **0.15 (0.06, 0.24)** | **<0.001** |
| Female Sex | **-3.01 (-4.21, -1.81)** | **<0.001** | - | - | - | - |
| BMI | **-0.22 (-0.34, -0.10)** | **<0.001** | **-0.19 (-0.37, -0.00)** | **0.048** | **-0.25 (-0.41, -0.08)** | **0.003** |
| Benzodiazepine | 1.52 (-1.18, 4.22) | 0.269 | -1.88 (-5.82, 2.05) | 0.348 | 3.63 (-0.13, 7.39) | 0.059 |
| CVD History | 0.40 (-1.16, 1.97) | 0.613 | 0.82 (-1.03, 2.66) | 0.387 | 0.02 (-2.70, 2.74) | 0.988 |
| COPD | -1.58 (-7.65, 4.49) | 0.610 | 0.35 (-7.33, 8.03) | 0.929 | -3.57 (-13.14, 5.99) | 0.464 |
| MAP | -0.01 (-0.06, 0.03) | 0.597 | -0.05 (-0.11, 0.01) | 0.093 | 0.02 (-0.05, 0.10) | 0.526 |
| AHI | **-0.27 (-0.31, -0.23)** | **<0.001** | **-0.28 (-0.32, -0.23)** | **<0.001** | **-0.26 (-0.33, -0.19)** | **<0.001** |

Coefficients ( $\beta$ ) and 95% confidence intervals (CI) are derived from linear regression models where AUC above baseline is the dependent variable, statistically significant results ( $p < 0.05$ ) indicate covariates that drive higher or lower AUC, adjusting for the other listed covariates. BMI, body mass index; AHI, apnea-hypopnea index; MAP, mean arterial pressure; CVD, cardiovascular disease; COPD, chronic obstructive pulmonary disease. "Gender (Female)" denotes a binary variable comparing females to the reference (male) group. $\beta$ represents the average change in AUC per 1-unit increase in the continuous covariate or the mean difference relative to the reference group for categorical variables. Values in bold indicate a significant association, $p < 0.05$ .

**Table S5.** Association of PT With Daytime Sleepiness (ESS ≥10) Across Cohorts.

| | $\beta$ | z | p | OR (95%CI) |
| --- | --- | --- | --- | --- |
| SHHS-Male | -0.013 | -0.713 | 0.476 | 0.987 (0.952-1.023) |
| SHHS-Female | -0.026 | -1.596 | 0.11 | 0.974 (0.944-1.006) |
| MESA-male | 0.0354 | 1.083 | 0.279 | 1.036 (0.972-1.105) |
| MESA-female | 0.008 | 0.383 | 0.701 | 1.008 (0.968-1.050) |
| MrOS | -0.0012 | -0.026 | 0.979 | 0.999 (0.913-1.092) |
| SOF | -0.013 | -0.713 | 0.476 | 0.987 (0.952-1.023) |

\*Logistic regression results are presented as the log-odds coefficient ( $\beta$ ), z-statistic (z), p-value, and odds ratio (OR) with 95% confidence interval. Models adjust for relevant demographic and clinical covariates in each cohort.

**Table S6.** Association of PT-NREM With Daytime Sleepiness (ESS ≥10) Across Cohorts.

| | $\beta$ | z | p | OR (95%CI) |
| --- | --- | --- | --- | --- |
| SHHS-Male | -0.0049 | -0.317 | 0.751 | 0.995 (0.966-1.026) |
| SHHS-Female | **-0.0287** | **-2.263** | **0.024** | **0.972 (0.948-0.996)** |
| MESA-male | -0.0022 | -0.075 | 0.94 | 0.998 (0.941-1.057) |
| MESA-female | 0.0323 | 1.748 | 0.08 | 1.033 (0.996-1.071) |
| MrOS | 0.0165 | 0.427 | 0.67 | 1.017 (0.942-1.097) |
| SOF | -0.0049 | -0.317 | 0.751 | 0.995 (0.966-1.026) |

Logistic regression results are presented as the log-odds coefficient ( $\beta$ ), z-statistic (z), p-value, and odds ratio (OR) with 95% confidence interval. Models adjust for relevant demographic and clinical covariates in each cohort.

**Table S7.** Association of PT-REM With Daytime Sleepiness (ESS ≥10) Across Cohorts.

| | $\beta$ | z | p | OR (95%CI) |
| --- | --- | --- | --- | --- |
| SHHS-Male | 0.0183 | 1.802 | 0.072 | 1.018 (0.998-1.039) |
| SHHS-Female | -0.0082 | -0.734 | 0.463 | 0.992 (0.970-1.014) |
| MESA-male | -0.0028 | -0.12 | 0.904 | 0.997 (0.953-1.044) |
| MESA-female | -0.0106 | -0.751 | 0.452 | 0.989 (0.962-1.017) |
| MrOS | **0.084** | **2.778** | **0.005** | **1.088 (1.025-1.154)** |
| **SOF** | 0.0183 | 1.802 | 0.072 | 1.018 (0.998-1.039) |

Logistic regression results are presented as the log-odds coefficient ( $\beta$ ), z-statistic (z), p-value, and odds ratio (OR) with 95% confidence interval. Models adjust for relevant demographic and clinical covariates in each cohort.

**Table S8.** PT Quartiles and Mortality Risk Across Cohorts.

|  | Quartile | All-cause mortality |  | CVD-mortality |  |
| --- | --- | --- | --- | --- | --- |
|  |  | HR (95%CI) | P-value | HR (95%CI) | P-value |
| SHHS-Male | Q1 (<4.5 s) | 1.0 | - | 1.0 | - |
|  | Q2 (4.5 – 5.5 s) | 1.13 (0.89-1.44) | 0.321 | 0.91 (0.53-1.57) | 0.741 |
|  | Q3 (5.5-7.5 s) | 0.90 (0.72-1.13) | 0.370 | 1.15 (0.75-1.76) | 0.529 |
|  | Q4 (>7.5 s) | 1.13 (0.89-1.43) | 0.308 | 1.32 (0.85-2.04) | 0.222 |
| SHHS-Female | Q1 (<4.5 s) | 1.0 |  | 1.0 |  |
|  | Q2 (4.5 – 5.5 s) | 1.00 (0.76-1.32) | 0.993 | 1.18 (0.69-2.02) | 0.547 |
|  | Q3 (5.5-7.5 s) | 1.00 (0.78-1.29) | 0.975 | 1.36 (0.84-2.20) | 0.210 |
|  | Q4 (>7.5 s) | **1.32 (1.02-1.71)** | **0.036** | **1.75 (1.08-2.83)** | **0.023** |
| MESA-Male | Q1 (<4.5 s) | 1.0 |  | 1.0 |  |
|  | Q2 (4.5 – 5.5 s) | 1.31 (0.73-2.33) | 0.366 | 0.75 (0.24-2.40) | 0.632 |
|  | Q3 (5.5-7.5 s) | 1.26 (0.78-2.03) | 0.348 | 1.38 (0.65-2.96) | 0.401 |
|  | Q4 (>7.5 s) | 1.59 (0.98-2.59) | 0.062 | 1.03 (0.41-2.60) | 0.956 |
| MESA-Female | Q1 (<4.5 s) | 1.0 |  | 1.0 |  |
|  | Q2 (4.5 – 5.5 s) | 0.93 (0.48-1.83) | 0.841 | 0.37 (0.06-2.27) | 0.282 |
|  | Q3 (5.5-7.5 s) | 0.68 (0.36-1.28) | 0.228 | 0.37 (0.08-1.68) | 0.198 |
|  | Q4 (>7.5 s) | 1.13 (0.67-1.92) | 0.640 | 0.96 (0.42-2.20) | 0.918 |
| SOF-Female | Q1 (<4.5 s) | 1.0 |  | 1.0 |  |
|  | Q2 (4.5 – 5.5 s) | 0.70 (0.33-1.51) | 0.367 | 0.34 (0.07-1.59) | 0.169 |
|  | Q3 (5.5-7.5 s) | 1.63 (0.94-2.82) | 0.079 | 1.88 (0.86-4.11) | 0.114 |
|  | Q4 (>7.5 s) | 1.33 (0.75-2.34) | 0.325 | 1.23 (0.53-2.87) | 0.626 |
| MrOS-Male | Q1 (<4.5 s) | 1.0 |  | 1.0 |  |
|  | Q2 (4.5 – 5.5 s) | 0.96 (0.79-1.18) | 0.702 | 1.13 (0.70-1.84) | 0.609 |
|  | Q3 (5.5-7.5 s) | 1.04 (0.87-1.25) | 0.644 | 1.10 (0.70-1.71) | 0.689 |
|  | Q4 (>7.5 s) | **1.31 (1.09-1.57)** | **0.004** | **1.69 (1.09-2.60)** | **0.018** |

Values represent hazard ratios (HRs) and 95% confidence intervals (CIs) for all-cause and cardiovascular disease (CVD) mortality across quartiles of Peak Time (PT), stratified by sex and cohort. Quartile 1 (<4.5 s) serves as the reference group. PT was modeled as a categorical variable in Cox proportional hazards regression, fully adjusted for covariates. Bold values indicate statistical significance ( $p < 0.05$ ) based on the Wald test.

**Table S9.**
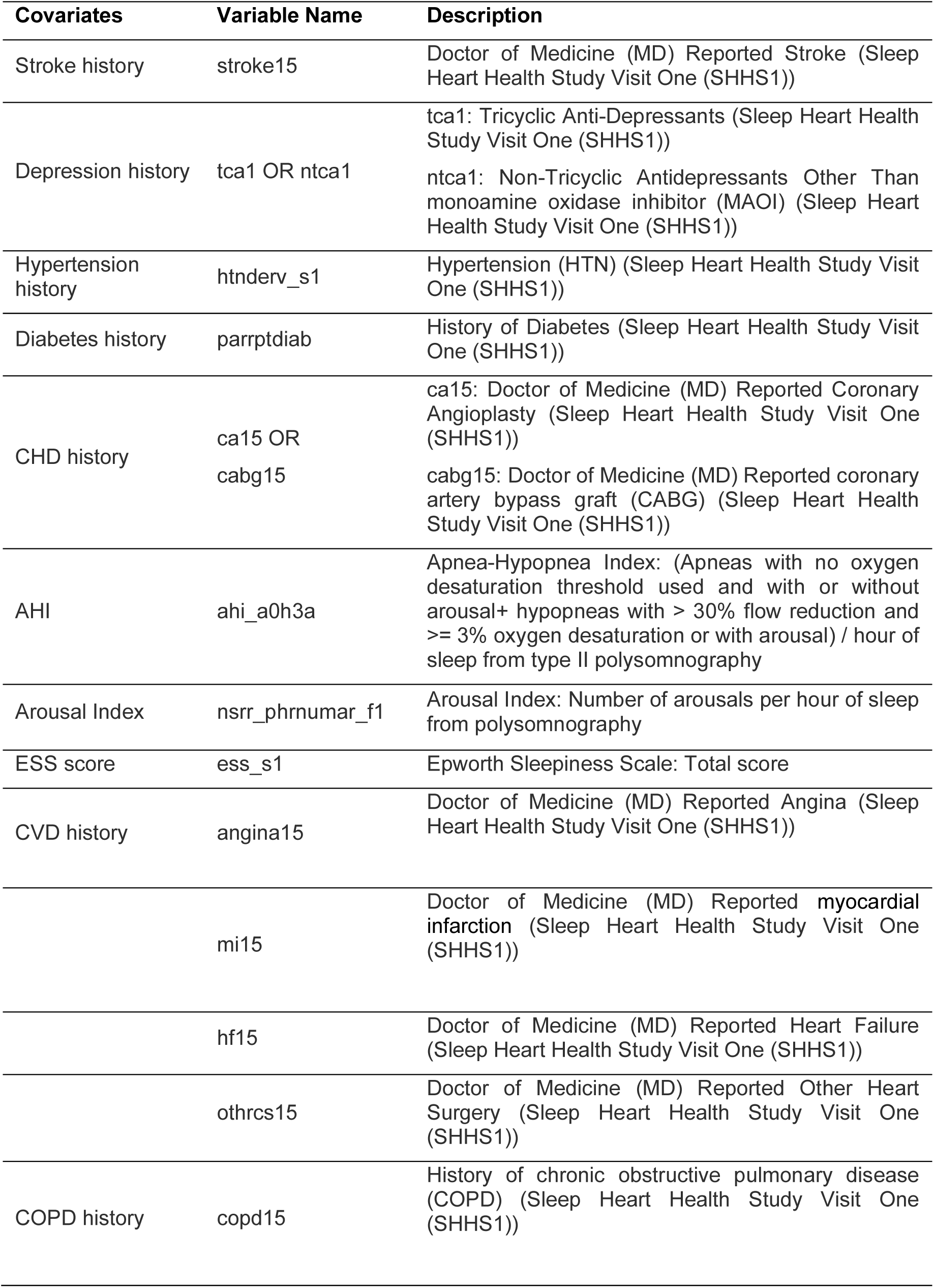
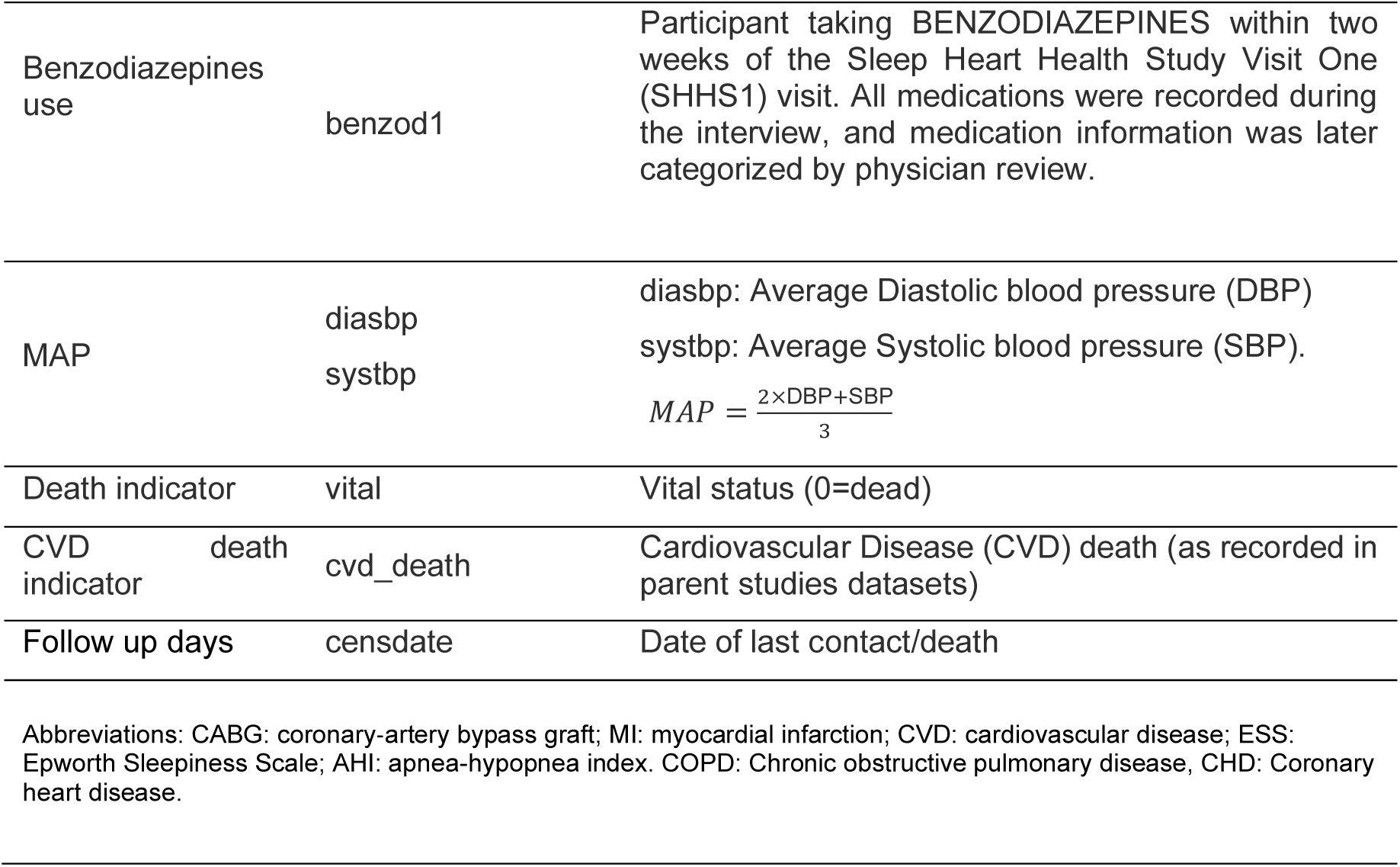
Definition of Covariates in SHHS.

**Table S10.**
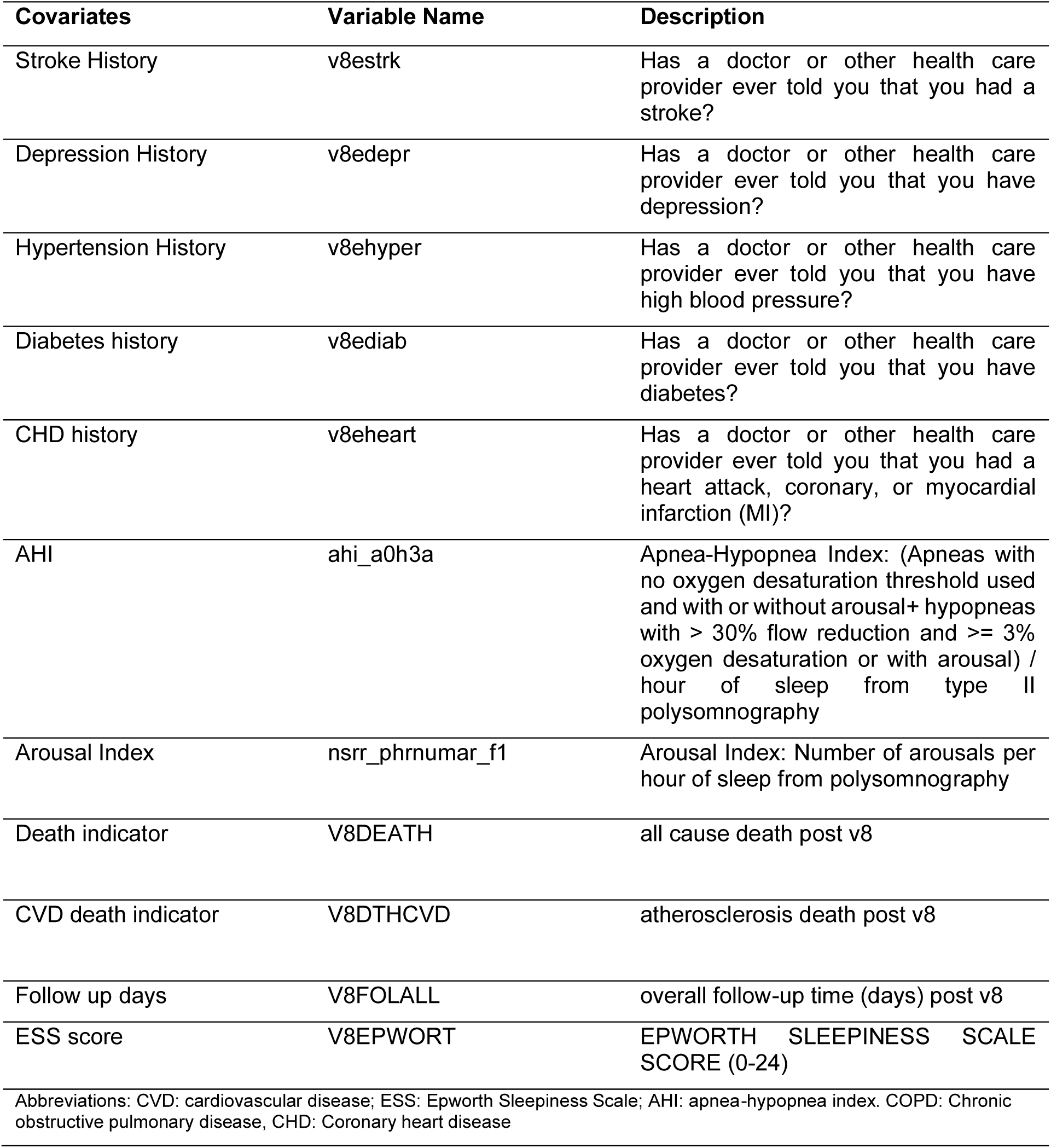
Definition of covariates in SOF.

**Table S11.**
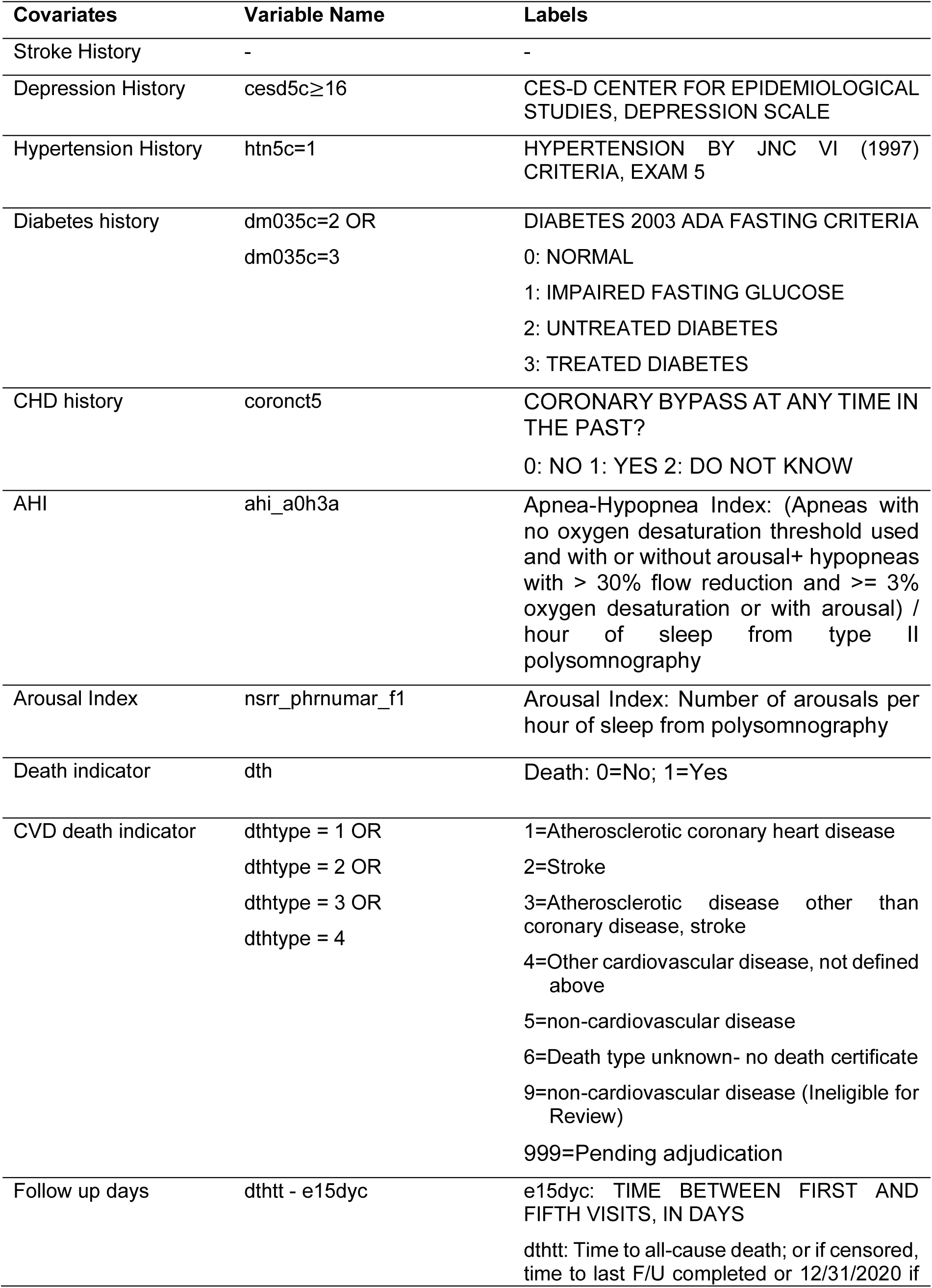
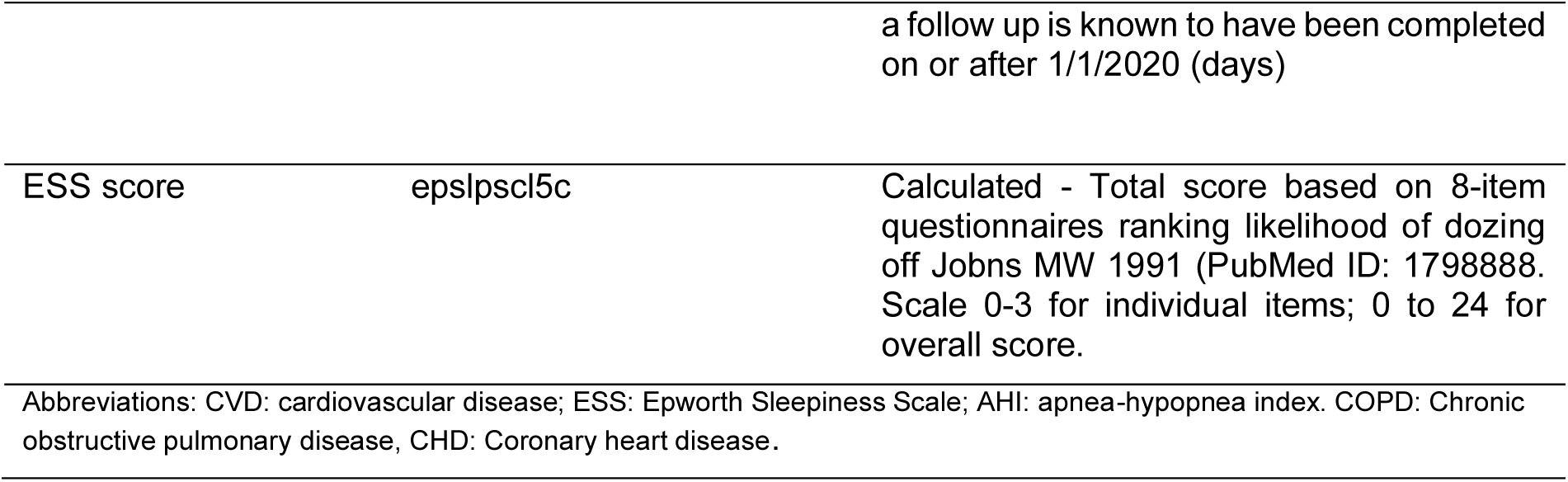
Definition of Covariates in MESA.

**Table S12.**
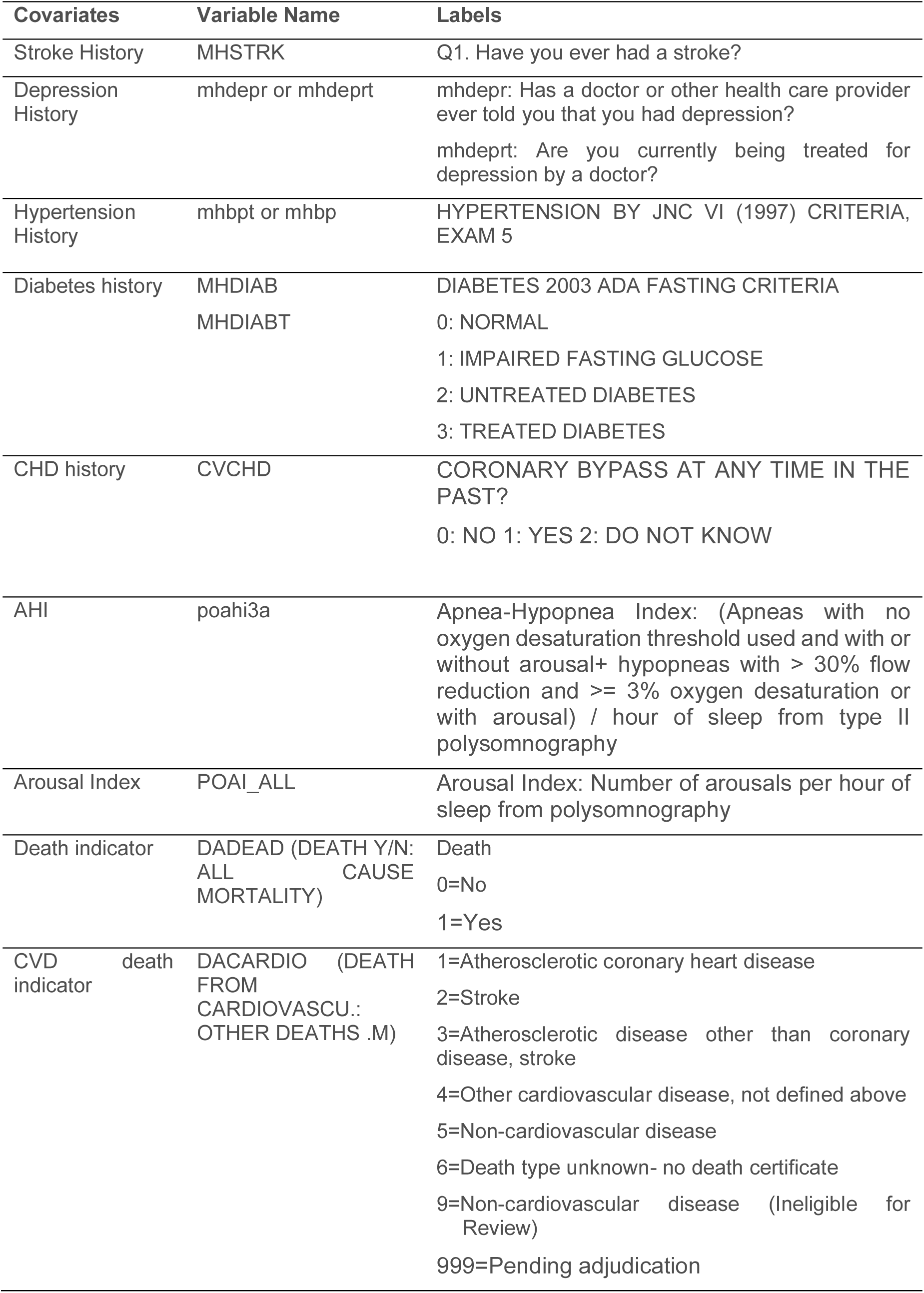
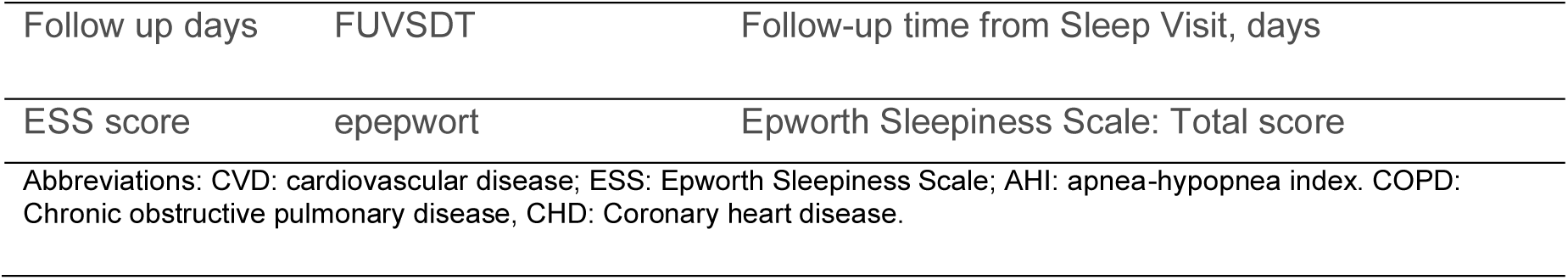
Definition of Covariates in MrOS.

